# Genetic subtyping of obesity reveals biological insights into the uncoupling of adiposity from its cardiometabolic comorbidities

**DOI:** 10.1101/2025.02.25.25322830

**Authors:** Nathalie Chami, Zhe Wang, Victor Svenstrup, Virginia Diez Obrero, Daiane Hemerich, Yi Huang, Hesam Dashti, Eleonora Manitta, Michael H. Preuss, Kari E. North, Louise Aas Holm, Cilius E Fonvig, Jens-Christian Holm, Torben Hansen, Camilla Scheele, Alexander Rauch, Roelof A.J. Smit, Melina Claussnitzer, Ruth J.F. Loos

## Abstract

Obesity is a highly heterogeneous disease that cannot be captured by one single adiposity trait. Here, we performed a multi-trait analysis to study obesity in the context of its common cardiometabolic comorbidities, acknowledging that not all individuals with obesity suffer from cardiometabolic comorbidities and that not all those with normal weight clinically present without them. We leveraged individual-level genotype-phenotype data of 452,768 individuals from the UK Biobank and designed uncoupling phenotypes that are continuous and range from high adiposity with a healthy cardiometabolic profile to low adiposity with an unhealthy cardiometabolic profile. Genome-wide association analyses of these uncoupling phenotypes identified 266 independent variants across 205 genomic loci where the adiposity-increasing allele is also associated with a lower cardiometabolic risk. Consistent with the individual variant effects, a genetic score (GRS_uncoupling_) that aggregates the uncoupling effects of the 266 variants was associated with lower risk of cardiometabolic disorders, including dyslipidemias (OR=0.92, P=1.4×10^-89^), type 2 diabetes (OR=0.94, P=6×10^-21^), and ischemic heart disease (OR=0.96, P=7×10^-11^), despite a higher risk of obesity (OR=1.16, P=4×10^-108^), which is in sharp contrast to the association profile observed for the adiposity score (GRS_BFP_). Nevertheless, a higher GRS_uncoupling_ score was also associated with a higher risk of other, mostly weight-bearing disorders, to the same extent as the GRS_BFP_. The 266 variants clustered into eight subsets, each representing a genetic subtype of obesity with a distinct cardiometabolic risk profile, characterized by specific underlying pathways. Association of GRS_uncoupling_ and GRS_BFP_ with levels of 2,920 proteins in plasma found 208 proteins to be associated with both scores. The majority (85%) of these overlapping GRS-protein associations were directionally consistent, suggesting adiposity-driven effects. In contrast, levels of 32 (15%) proteins (e.g. IGFBP1, IGFBP2, LDLR, SHBG, MSTN) had opposite directional effects between GRS_BFP_ and GRS_uncoupling_, suggesting that cardiometabolic health, and not adiposity, associated with their levels. Follow-up analyses provide further support for adipose tissue expandability, insulin secretion and beta-cell function, beiging of white adipose tissue, inflammation and fibrosis. They also highlight mechanisms not previously implicated in uncoupling, such as hepatic lipid accumulation, hepatic control of glucose homeostasis, and skeletal muscle growth and function. Taken together, our findings contribute new insights into the mechanisms that uncouple adiposity from its cardiometabolic comorbidities and illuminate some of the heterogeneity of obesity, which is critical for advancing precision medicine.

## INTRODUCTION

Obesity is a major risk factor for a variety of cardiometabolic disease outcomes and is the consequence of intricate interactions between genes and environment^1–4^. While an increasingly obesogenic environment is a key driver of the current obesity epidemic, people’s innate genetic predisposition determines the extent of weight gain in this environment. Genome-wide association studies (GWAS) have identified more than 1,000 genetic loci associated with obesity risk^5^. Subsequent tissue enrichment analyses have pointed to the central nervous system as playing a key role in body weight regulation^5,6^. Despite these insights into the overarching biology, our understanding of the mechanisms that control body weight is still limited. This could be, in part, because GWASs have so far considered obesity as a homogeneous condition, with each analysis focusing on one adiposity trait at a time, most often body mass index (BMI). Such single-trait GWASs ignore the vast heterogeneity among individuals with obesity in, for example, etiology, life course trajectory and cardiometabolic comorbidities.

Therefore, while single-trait GWASs have identified numerous loci^5^, they likely represent only a subset of the mechanisms underlying obesity. Multi-trait GWASs, on the other hand, hold the potential to reveal more layers of the underlying biology. For example, we and others have performed genome-wide searches for loci that uncouple excess adiposity from cardiometabolic risk and identified 87 loci of which the adiposity-increasing allele associates with lower cardiometabolic risk^7–14^. Pathway and tissue enrichment analyses for such uncoupling loci have implicated peripheral mechanisms, such as fat distribution, adipocyte function and differentiation, and inflammation, but not the central nervous system^15^.

Here, we build upon our previous work^11^ and leverage individual-level data from 452,768 European participants in the UK Biobank to perform a comprehensive multi-trait genome-wide screen. We aimed to identify novel genetic loci that uncouple adiposity from cardiometabolic comorbidities by analyzing three adiposity and eight cardiometabolic traits, including lipid, glycemic, and blood pressure traits. We identified 205 genetic loci, harboring 266 lead variants, where the adiposity-increasing allele is associated with a lower risk of at least one cardiometabolic trait. Follow-up analyses included genetic clustering, genetic score construction, tissue and pathway enrichment, gene prioritization, phenome-wide association studies (PheWAS), and proteomic profiling and led to the identification of genetic subtypes of obesity with distinct cardiometabolic risk profiles and their association with specific pathways and serum protein profiles.

## RESULTS

### Genome-wide screen identifies 266 adiposity-increasing alleles with protective effects on cardiometabolic health

We performed a genome-wide screen in up to 452,768 individuals of European ancestry from the UK Biobank to identify adiposity increasing loci that have protective effects on cardiometabolic health. We analyzed three adiposity traits (body mass index (BMI), body fat percentage (BFP) and waist-to-hip ratio (WHR)) and eight cardiometabolic traits (total cholesterol (TC), LDL cholesterol (LDL-C), HDL cholesterol (HDL-C), triglycerides (TG), random glucose, and HbA1c levels, and systolic (SBP) and diastolic blood pressure (DBP)) (**Methods, Fig. 1 and Supplementary Table 1).** We first created 24 “bi-traits”, which combine an adiposity and a cardiometabolic trait into a new phenotype, obtained by subtracting standardized values of one of the eight cardiometabolic traits from standardized values of one of the three adiposity traits. High values for a bi-trait represent high adiposity but low levels of the cardiometabolic trait and vice versa (**Methods and Extended Data** Fig. 1). We next performed GWAS for each of the 24 bi-traits and each of the 11 single traits from which the bi-traits were derived. Variants associated with bi-traits at genome-wide significance (P < 5×10^-10^), and for which the associations with both corresponding single traits reached nominal significance (P<10^-4^) and that also colocalized were considered uncoupling variants. As such, we identified 266 unique lead variants located in 205 loci (more than 1 Mb apart) across the 24 bi-traits (**Methods; Supplementary Tables 2-4).** Of the 205 uncoupling loci we identified, 139 have not previously been reported in the context of cardiometabolic uncoupling, whereas the remaining 66 replicate the majority of the 87 loci reported across earlier studies^7–15^. Twenty-one loci reported previously did not reach significance in our study because of the use of a more stringent significance threshold and differences in study design, traits and disease outcomes studied. Altogether, our analyses identified more than double the number of previously reported loci, which is in part driven by a larger sample size, but also by the use of individual-level data that allowed us to design new, continuous uncoupling phenotypes, as opposed to summary statistics used in previous studies^7–11,14^ **(Supplementary Tables 2 and 5).**

**Fig. 1:**
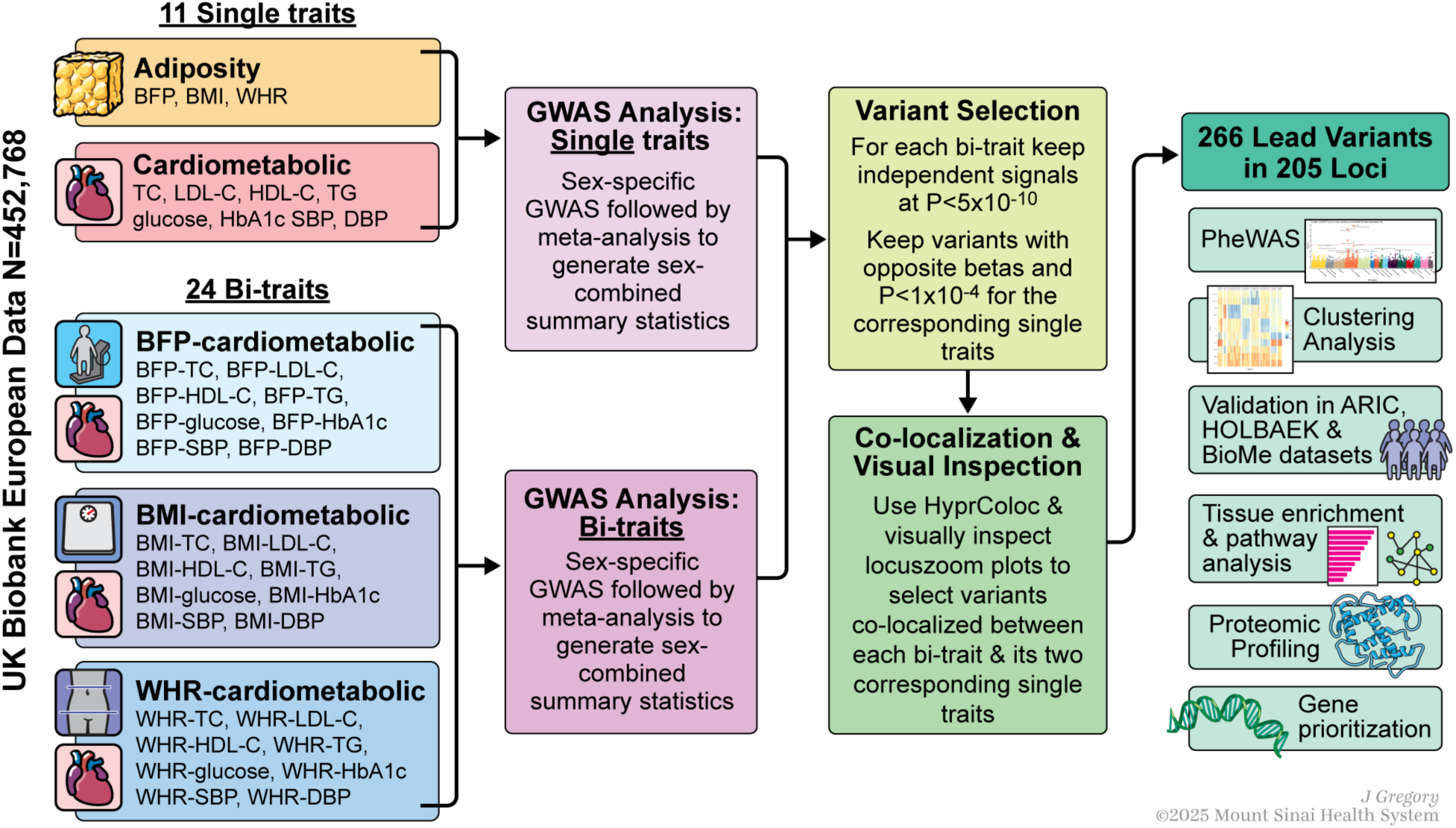
Study overview. Overall steps and traits analyzed in the study. Bi-traits are obtained by subtracting standardized values of a cardiometabolic trait from an adiposity trait. BMI: body mass index; BFP: body fat percentage; WHR: waist-to-hip ratio; TC: total cholesterol; LDL-C: LDL cholesterol; HDL-C: HDL cholesterol; TG: triglycerides; SBP: systolic blood pressure; DBP: diastolic blood pressure.

### A genetic risk score (GRS_uncoupling_) associated with higher adiposity, but lower cardiometabolic traits

To assess the aggregate effect of the uncoupling loci, we created a genetic risk score (GRS) comprised of the 266 variants, GRS_uncoupling_. To compare GRS_uncoupling_ with a proxy for adiposity that captures overall body fat without factoring in the cardiometabolic component, we created another GRS comprised of the 647 variants that were significantly (P<5×10^-10^) associated with BFP, GRS_BFP_ **(Methods)**. We tested the association of both GRSs with 13 adiposity traits and eight cardiometabolic traits **(Methods; Fig. 2A)**. Both GRSs were associated with higher values of most adiposity traits; with effect sizes for GRS_BFP_ tending to be generally larger than those for GRS_uncoupling_. However, compared to the GRS_BFP_, the GRS_uncoupling_ was associated with a more favorable fat distribution. Specifically, a higher GRS_uncoupling_ was associated with a lower MRI-derived visceral to abdominal subcutaneous adipose tissue volume (VAT/ASAT) ratio and no association with WHR, whereas a higher GRS_BFP_ was associated with a higher WHR, but not with VAT/ASAT ratio. Consistently, the GRS_uncoupling_ had a larger effect on gynoid than on android fat percentage, whereas the opposite was seen for the GRS_BFP_. Furthermore, a higher GRS_uncoupling_ was associated with less liver fat, whereas a higher GRS_BFP_ was associated with more liver fat.

**Fig. 2:**
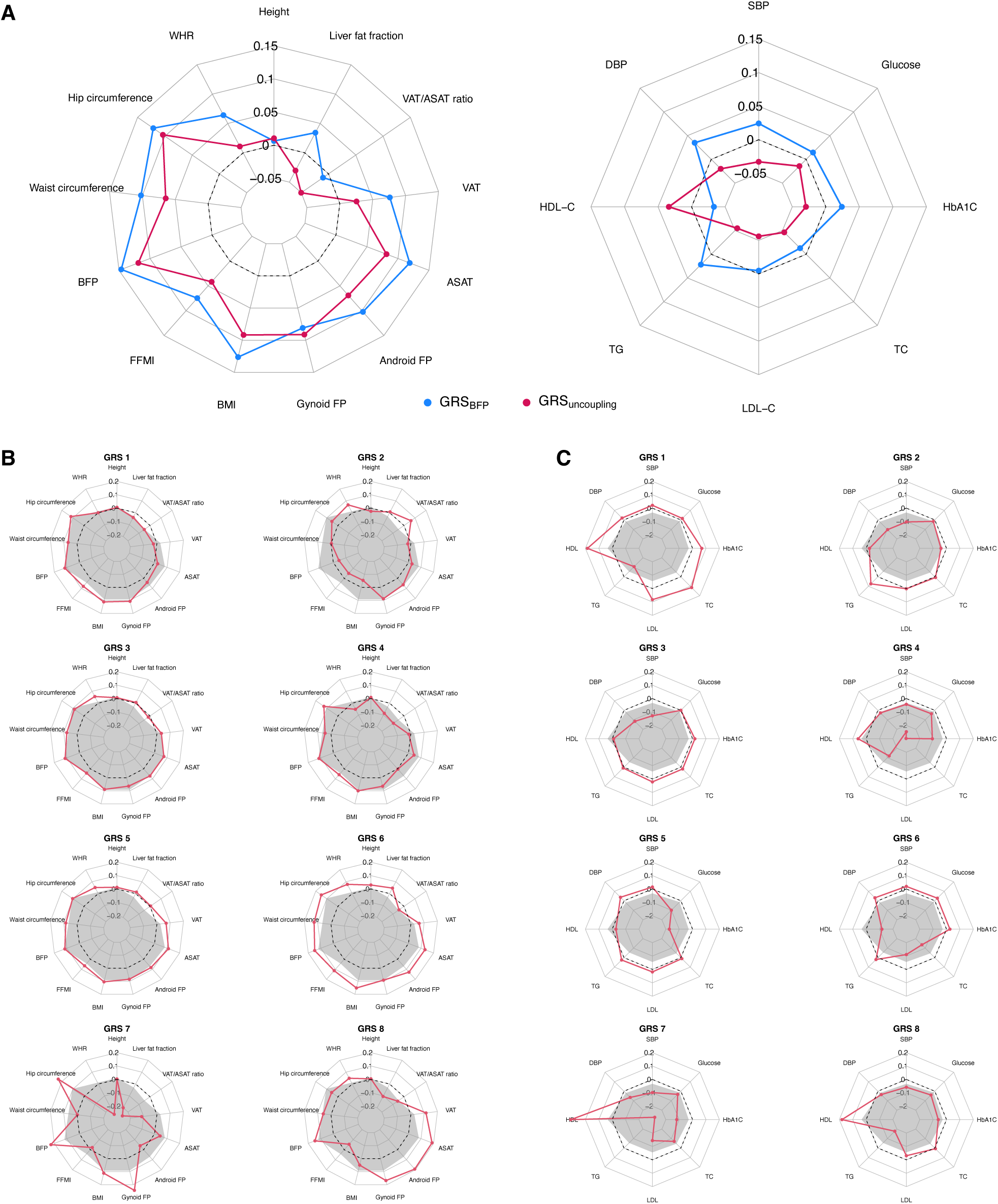
Associations of genetic risk scores with anthropometric and cardiometabolic traits in the UK Biobank. A. Estimated per 10 allele change effect sizes of GRS–trait associations in UK Biobank European ancestry population for GRS_uncoupling_ (in magenta) and GRS_BFP_ (in blue). B-C. Estimated per 10 allele change effect sizes of GRS–trait associations in UK Biobank European ancestry population for each cluster specific GRS (GRS 1-8, in red) and GRS_BFP_ (in gray). Dashed circles indicate Beta=0, indicating no association between each GRS and the trait. Points outside the circle represent positive GRS–trait associations, while those inside represent negative associations. The effect and reference alleles of GRS_2_, a cluster associated with lower WHR and higher blood pressure, were flipped in order to reflect a profile of higher adiposity and facilitate comparison with the other clusters.

For cardiometabolic traits, a higher GRS_uncoupling_ was associated with a healthier profile; i.e. lower levels of LDL-C, TC, TG, HbA1c, glucose, SBP and DBP and higher HDL-C. A higher GRS_BFP_, on the other hand, was associated with an unhealthy cardiometabolic profile; i.e. higher levels of TG, HbA1c, glucose, SBP, and DBP and lower HDL-C, but no effect on LDL-C or TC **(Fig. 2A and Supplementary Table 6**). This association signature of the GRS_uncoupling_ was significantly different from that of the GRS_BFP_ (P < 0.0001) **(Fig. 2A and Supplementary Table 6).**

In men and women separately, the association of GRS_uncoupling_ with adiposity traits tended to differ for the fat distribution traits. Specifically, the association with a more favorable fat distribution (WHR, VAT/ASAT) was more pronounced in women than in men (**Extended Data** Fig. 2). This was mostly due to a smaller effect on abdominal fat accumulation (waist circumference, android FP) in women compared to men, whereas the effect on peripheral fat accumulation (hip circumference, gynoid FP) was not substantially different between sexes. Despite these differences in fat distribution, associations with cardiometabolic traits were similar in men and women. No sex-specific effects for either anthropometric or cardiometabolic traits were observed for the GRS_BFP_ **(Extended Data** Fig. 2).

### GRS_uncoupling_ associates with protective effect on cardiometabolic outcomes but with increased risk of weight-bearing diseases

To better understand the clinical impact of genetic predisposition to adiposity and cardiometabolic comorbidities, we performed a phenome-wide association analysis (PheWAS) between the two GRSs (GRS_uncoupling_, GRS_BFP_) and 10,965 disease outcomes in the UK Biobank (N = 373,747) **(Methods, Supplementary Table 7, Fig. 3).** While GRS_uncoupling_ is associated with a healthier cardiometabolic profile, it is also associated with an increased risk for a wide range of other diseases, often weight-bearing diseases, to the same extent as GRS_BFP_. Specifically, a higher GRS_uncoupling_ was associated with a lower risk of conditions related to lipoprotein metabolism (OR=0.92, P=1.4×10^-89^), essential primary hypertension (OR=0.96, P=1.7×10^-27^), non-insulin dependent diabetes (OR=0.94, P=5.6×10^-21^), ischaemic heart disease (OR=0.96, P=7.4×10^-11^), angina (OR=0.96, P=1.6×10^-8^), and acute myocardial infarction (OR=0.96, P=3.6×10^-6^) **(Fig. 3),** whereas a higher GRS_BFP_ was associated with increased risk of these conditions. However, a higher GRS_uncoupling_ was associated with an increased risk of non-cardiometabolic, weight-related conditions, including cellulitis (OR=1.05, P=1.14×10^-11^), gonarthrosis (OR=1.06, P=9.9 x 10^-25^), and varicose veins (OR=1.08, P=7.9×10^-26^), similar to GRS_BFP_ **(Fig. 3)**.

**Fig. 3:**
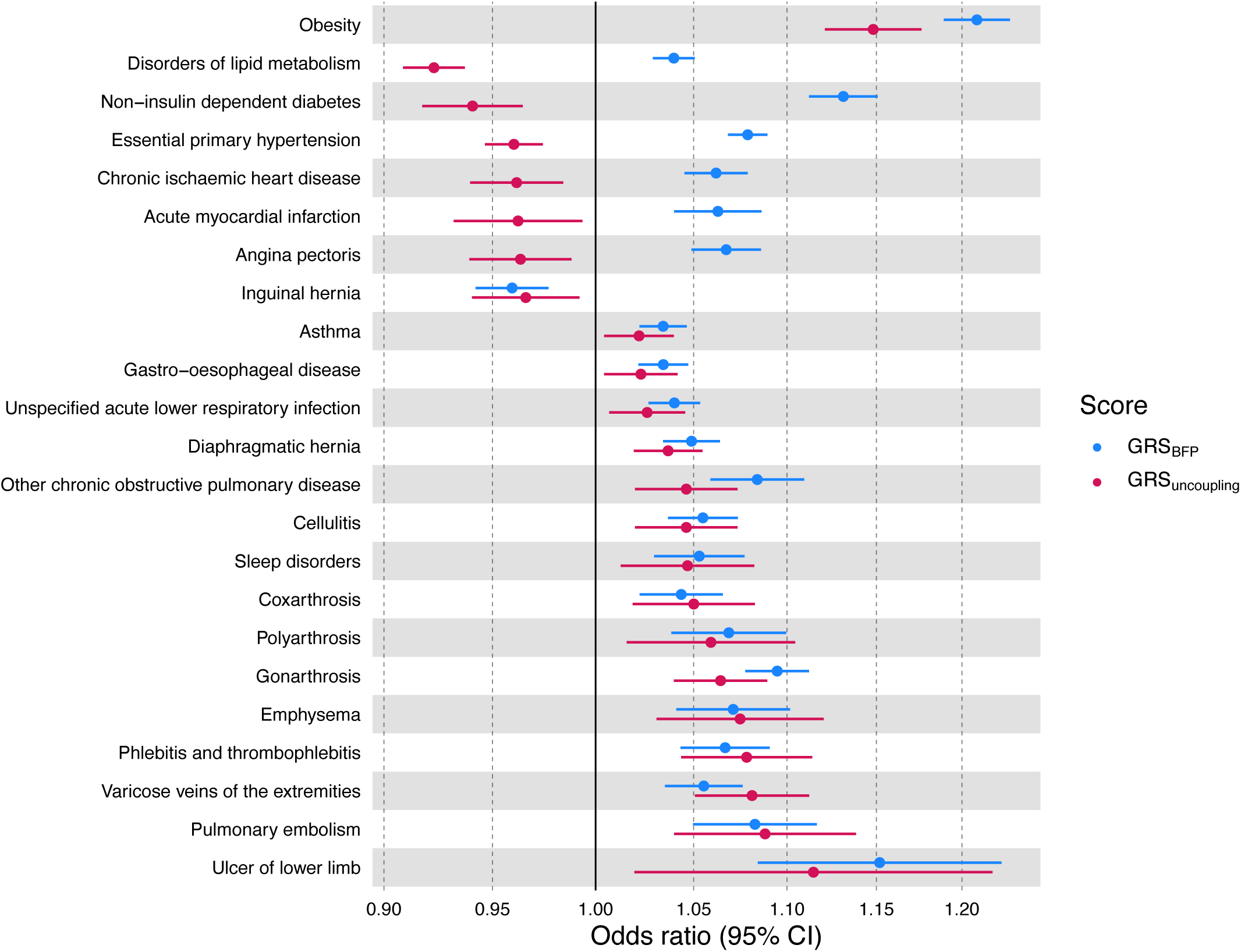
Association of genetic risk scores with disease outcomes in the UK Biobank. Phenome-wide association results of disease outcomes and GRS_BFP_ (in blue) and GRS_uncoupling_ (in magenta) performed using PHESANT in the European population. Odds ratios represent effect size estimates per 10 risk-allele increments.

### The 266 uncoupling lead variants group into 8 clusters with distinct cardiometabolic signatures

Even though each of the 266 uncoupling SNPs are associated with increased adiposity, and have a protective cardiometabolic effect, their association signatures vary substantially. We sought to identify genetically defined subgroups based on similarity of association with adiposity and cardiometabolic traits using NavMix clustering analyses **(Methods).** As such, we identified eight clusters with distinct association signatures **(Fig. 4).** Specifically, variants in three clusters (4,7 and 8) were associated with increased adiposity and an overall healthier cardiometabolic profile, with protective effects across multiple trait groups, whereas lead variants in the other five clusters were associated with increased adiposity with protective effects on only one of the cardiometabolic trait groups.

**Fig. 4:**
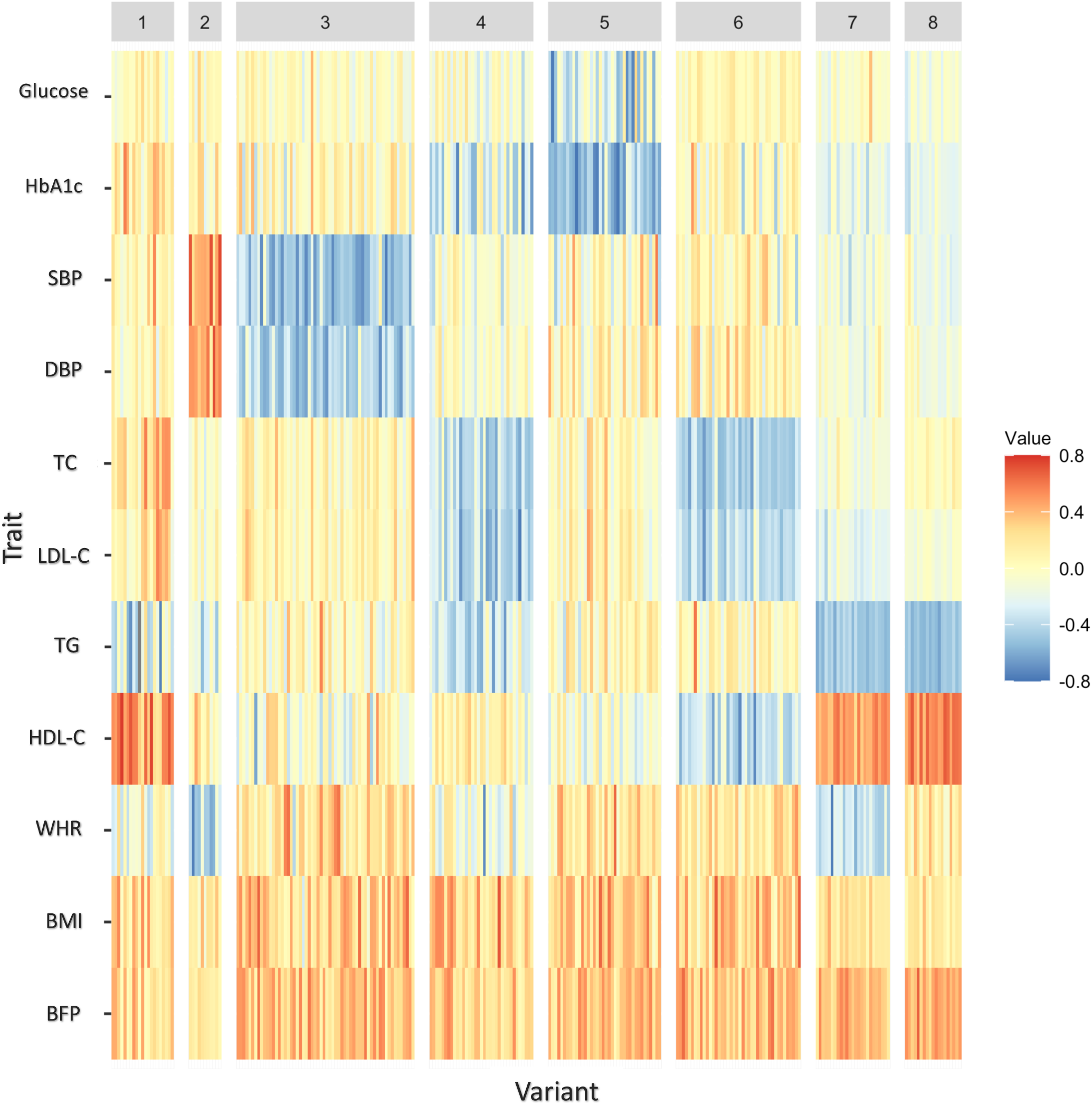
Heatmap of the association of the lead variants with adiposity and cardiometabolic traits. Clustering of the 266 uncoupling lead variants using the NAvMix algorithm identified eight clusters. The color coding represents beta values ranging from negative (blue) to positive (red). Associations are expressed by the BFP-increasing allele as the effect allele to enable comparison across traits.

We then calculated cluster-specific GRSs, GRS_1_-GRS_8,_ which aggregate the effects of lead variants in each cluster and tested their association with anthropometric and cardiometabolic traits (**Methods**). These cluster-specific GRSs displayed a distinct association signature, consistent with the general associations observed for the individual lead variants in each cluster **(Methods, Fig.2 B-C and Supplementary Table 6).** For example, GRS_4_, GRS_7_, and GRS_8_ are associated with two or more cardiometabolic trait groups. Specifically, GRS_4_ is associated with higher overall adiposity (higher BFP, BMI, fat free mass index (FFMI)), and favorable lipid and glycemic profiles (lower LDL-C, TC, TG, higher HDL-C and lower HbA1c), possibly mediated through a more favorable fat distribution (lower WHR, VAT/ASAT, and android FP). GRS_7_ and GRS_8_ were mainly associated with lower TG and higher HDL-C, lower HbA1c and blood pressure **(Figures 2B-C and Supplementary Table 6)**. Besides the stronger effects on HDL-C and TG, GRS_7_ differs from GRS_8_ in its association with adiposity. GRS_7_ has a strong effect on body fat distribution (e.g. lower WHR, VAT/ASAT and higher gynoid, lower android FP). On the other hand, GRS_8_ is associated with higher overall adiposity (BFP, BMI, gynoid & android FP, ASAT & SAT), lower fat free mass (FFMI), without an obvious effect on fat distribution **(Fig.2B and Supplementary Table 6)**. GRS_3_ and GRS_5_ are similarly associated with higher overall adiposity, with GRS_3_ being associated with lower blood pressure (SBP & DBP) and GRS_5_ with lower glycemic traits (glucose & HbA1c). GRS_1_ is associated with greater overall body size (BMI, FFMI, BFP), and with lower TG and higher HDL-C levels, but also with higher levels of LDL-C, TC, and HbA1c. Of all cluster-specific GRSs, GRS_6_ has the strongest effect on overall body size. On the cardiometabolic side, GRS_6_’s profile is opposite to that of GRS_1_; as GRS_6_ is associated with lower LDL-C and TC levels, but also with higher TG and lower HDL-C levels, and high blood pressure and glycemic traits **(Fig.2B-C and Fig. 4**). Finally, GRS_2_ is the only cluster where the adiposity effect is driven mainly by an association with WHR. A higher GRS_2_ is associated with higher fat gynoid accumulation (WHR, VAT/ASAT), but lower blood pressure. In terms of disease outcomes, we observed cluster-specific associations with disease outcomes consistent with the characteristics of each cluster **(Supplementary Table 7).**

These findings highlight the potential for GRSs to identify subgroups among individuals with obesity. The GRS_uncoupling_ quantifies people’s risk of obesity without cardiometabolic comorbidities, whereas the GRS_BFP_ quantifies people’s risk of obesity with cardiometabolic comorbidities. The cluster-specific GRSs provide further granularity to this subgroup identification.

### GRS_uncoupling_ association signature validated in an independent cohort

To validate our findings, we tested the association of GRS_uncoupling_, the 8 cluster-specific GRSs (GRS_1-8_) and GRS_BFP_ with adiposity and cardiometabolic traits in the ARIC study, a population-based cohort (N = 15,792 individuals) **(Methods).** Similar to the UK Biobank, in ARIC, GRS_uncoupling_ was associated with a healthier cardiometabolic profile with lower levels (Beta<0) for glucose, TC, LDL-C, TG, SBP and DBP and higher HDL compared to GRS_BFP_. Both GRSs were associated with high adiposity including BMI, waist and hip circumference, and WHR albeit GRS_BFP_ generally having higher effect sizes **(Fig. 5A and Supplementary Table 8).** Similarly, the association signature of the cluster-specific GRSs with adiposity and cardiometabolic traits corresponded to that of the UK Biobank **(Fig. 5B-C and Supplementary Table 8).**

**Fig. 5:**
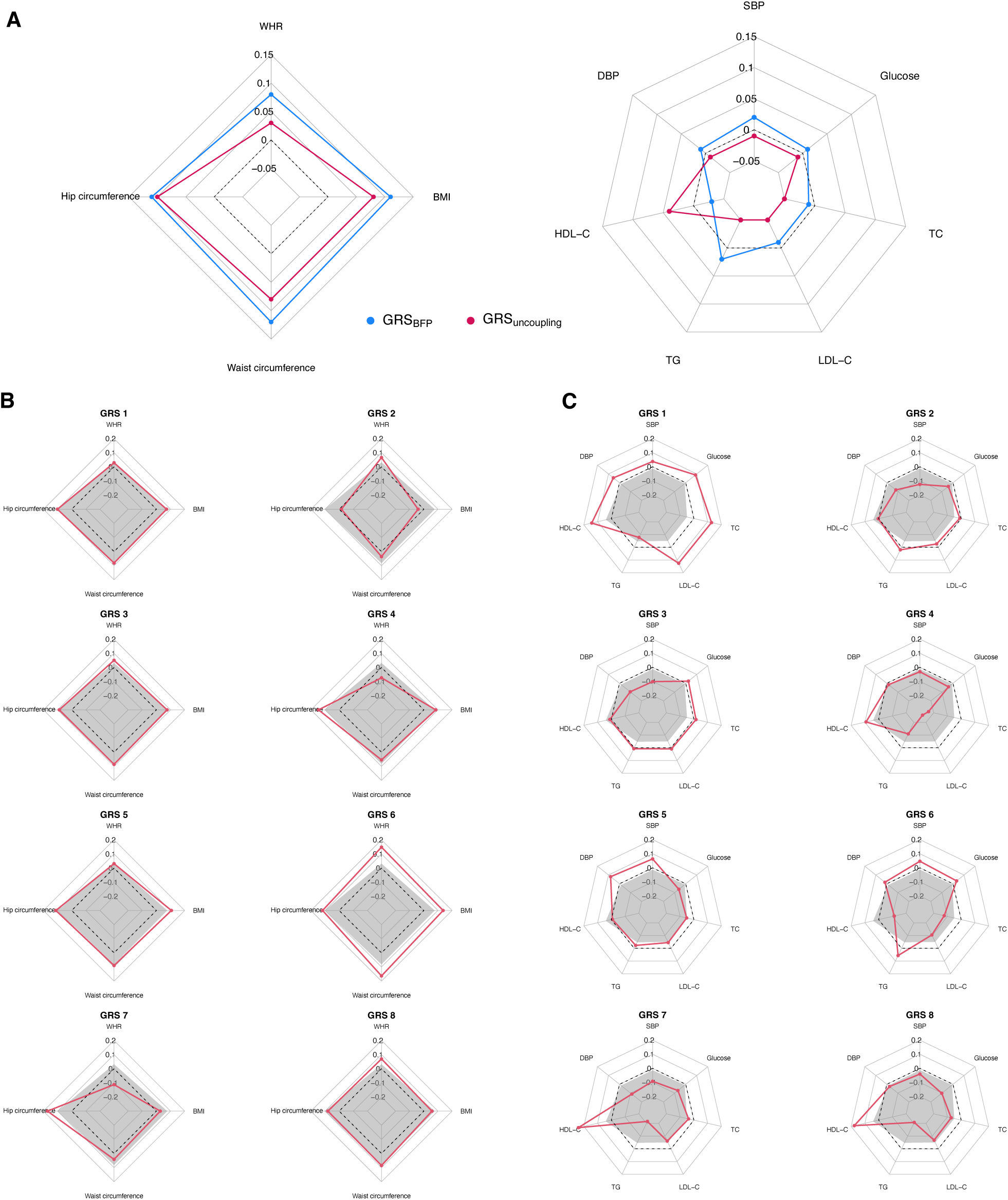
Associations of genetic risk scores with anthropometric and cardiometabolic traits in the ARIC study. **A.** Estimated per 10 allele change effect sizes of GRS–trait associations in 9,240 unrelated European ancestry participants of the ARIC study for GRS_uncoupling_ (in magenta) and GRS_BFP_ (in blue). B-C. Estimated per 10 allele change effect sizes of GRS–trait associations in UK Biobank European ancestry population for each cluster specific GRS (GRS 1-8, in red) and GRS_BFP_ (in gray). Dashed circles indicate Beta=0, indicating no association between each GRS and the trait. Points outside the circle represent positive GRS– trait associations, while those inside represent negative associations. The effect and reference alleles of GRS_2_, a cluster associated with lower WHR and higher blood pressure, were flipped in order to reflect a profile of higher adiposity and facilitate comparison with the other clusters.

### GRS_uncoupling_ is associated with lower T2D and CHD incidence

We also tested associations of GRS_uncoupling_ and GRS_BFP_ with incident coronary heart disease (CHD) and T2D in ARIC and the Mount Sinai Bio*Me* biobank, an electronic medical record linked biobank (N =50,000). A higher GRS_uncoupling_ was associated with a significantly lower risk of both incident CHD (hazard ratio [HR] 0.95; 95% CI 0.92 - 0.98, per 10 allele change in GRS) and T2D (HR 0.96; 95% CI 0.92 - 0.99) **(Supplementary Table 9)**, whereas a higher GRS_BFP_ was associated with higher risk of developing T2D (HR 1.04, 95% CI 1.01 - 1.07), but not CHD (HR 1.01, 95% CI 0.99 - 1.04).

### GRS_uncoupling_ already predisposes to higher adiposity but a more favorable cardiometabolic profile early in life

Among 3,457 Danish children and adolescents from the HOLBAEK study, the GRS_BFP_ and GRS_uncoupling_ were associated with higher overall adiposity **(Extended Data** Fig. 3A). Within the population-based cohort subset of 1,811 participants (**Methods**), both GRSs showed associations with BMI, to the same extend as those observed in the UK Biobank (e.g., GRS_uncoupling_: 0.09 in UK Biobank and 0.08 in HOLBAEK) **(Supplementary Table 10**). Participants with a higher GRS_BFP_ were more likely to present with a dysglycemic profile (higher HOMA-IR, insulin, C-peptide), whereas the GRS_uncoupling_ associated with a neutral glycemic profile and lower alkaline phospatase (ALP) levels **(Extended Data** Fig. 3B). Moreover, having a higher GRS_uncoupling_ associated with a lower risk of dyslipidemia (OR 0.89 (95%CI: 0.82, 0.97)) **(Extended Data** Fig. 3C).

### Uncoupling loci and overall adiposity loci have distinct tissue and pathway enrichment profiles

We next performed enrichment analyses for the 266 uncoupling lead variants, using DEPICT **(Methods)**, to identify the tissues and gene sets in which potential candidate genes may be acting and compared these with the results for the 647 BFP variants. Genes located in BFP-associated loci were mainly enriched in the central nervous system (CNS) (P = 0.002), consistent with previous observations for BMI-associated loci^6^. In contrast, genes located in the uncoupling loci were not enriched in the CNS (P=0.74) and were mostly enriched in peripheral tissues, including adipocytes and adipose tissue (P = 7×10^-7^), cardiovascular (P=1.4×10^-5^), digestive (P=7.8×10^-4^), endocrine (P=2.5×10^-4^), and musculoskeletal systems (P=2.5×10^-5^) **(Fig.6, Supplementary Table 11)**.

**Fig. 6:**
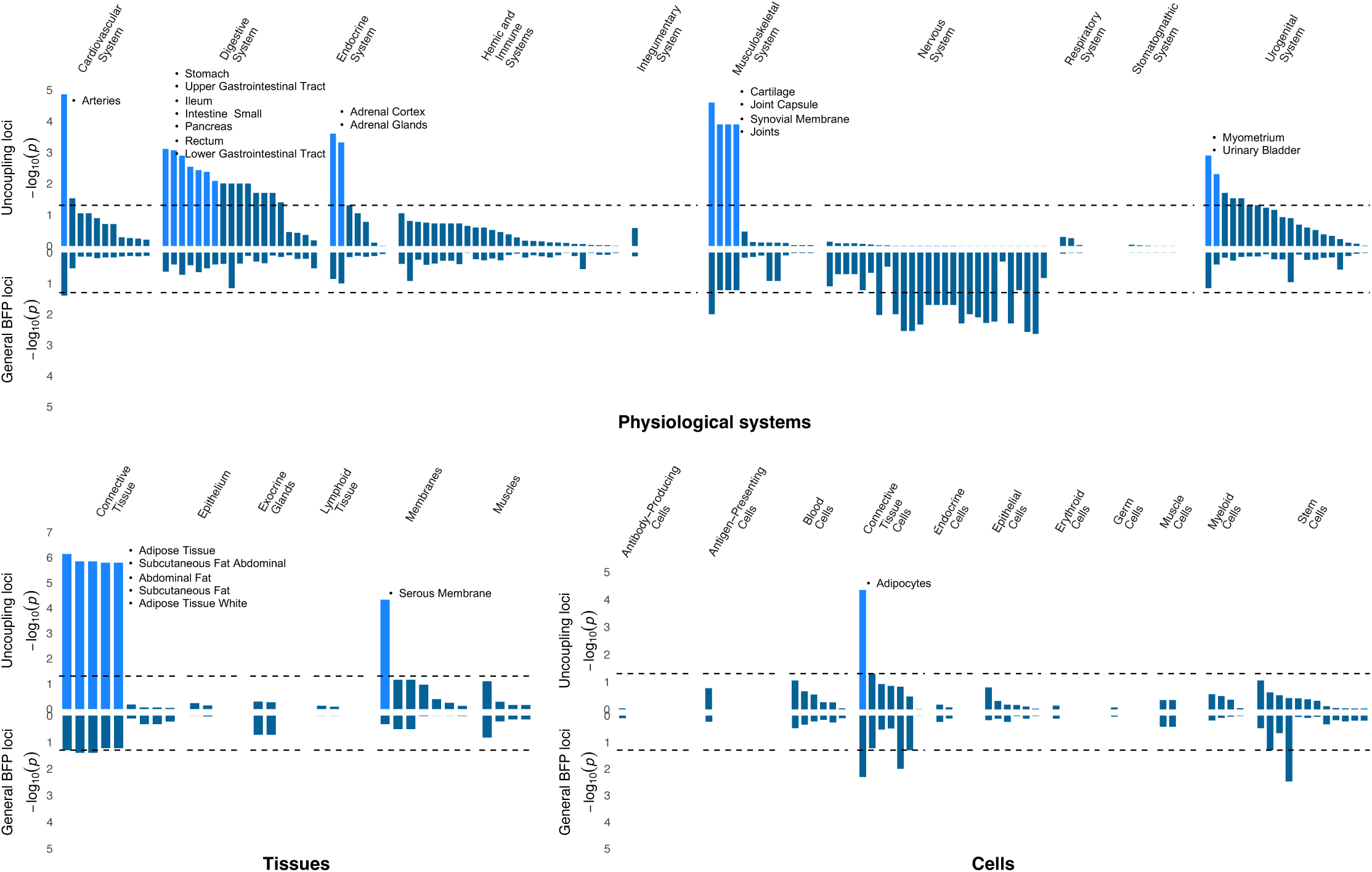
Physiological systems, tissue and cell-type enrichment analyses for uncoupling loci (top) and general body fat percentage loci (bottom). DEPICT results were based on summary statistics generated from the association analyses of the 266 lead SNPs with body fat percentage. We restricted our analyses to the Gene Ontology, KEGG, and REACTOME pathways terms. Results that passed the FDR corrected significance threshold (FDR < 0.05) are highlighted in bright blue. The dashed line represents the nominal significance level (P<0.05).

In gene set enrichment analyses for the uncoupling loci, we replicate previous findings for uncoupling loci, implicating insulin signaling, glucose homeostasis, lipid metabolism, immune and inflammatory response, and pathways related to adipose tissue biology (**Supplementary Table 12)**. In addition, we identify gene sets not previously implicated, including those related to vascular development, skeletal muscle development, liver development, kidney development, cartilage development, circadian rhythm, sex differentiation, and response to hypoxia, thereby implicating new biological processes.

On the other hand, genes in BFP loci were enriched for neurodevelopment and neuron differentiation including pathways related to brain development and regulatory mechanisms of the nervous system, consistent with previous literature^6^ **(Extended Data** Fig. 4 **and Supplementary Table 12).**

Cluster-specific gene set enrichment analyses implicated biological pathways that are consistent with their cardiometabolic health profile. For example, triglyceride lipase activity was highlighted for cluster 1, cardiovascular-related gene sets for cluster 2, muscle-related gene sets for cluster 4, transcriptional regulation of white adipose tissue regulation for cluster 7 **(Fig. 4 and Supplementary Table 12)**.

### The uncoupling GRSs are defined by distinct plasma proteomic profiles

To further characterize the biological signature of the GRSs, we assessed their association with 2,920 Olink-derived plasma proteins in the UK Biobank (**Methods and Supplementary Tables 13-14**). In line with the finding that 80% of these proteins are associated with measured BMI in this population^16^, both the GRS_BFP_ and GRS_uncoupling_ were associated (FDR<0.005) with a substantial number of protein levels: 915 (i.e., 31.3%) and 337 (11.5%), respectively, of which 208 protein were associated with both GRSs. Of these 208 overlapping proteins, 176 (85%) showed directional consistency in effect estimates, likely reflecting primarily adiposity-driven effects (**Extended Data** Fig. 5 and **Supplementary Table 13**). Notable examples include proteins shown to associate most strongly with higher BMI^16^, such as leptin (LEP), fatty acid binding protein 4 (FABP4), and pro-adrenomedullin (ADM). On the other hand, 32 proteins showed directionally opposing effects, potentially highlighting health-driven effects (**Fig.7**). These include proteins involved in lipid transport (e.g. LDLR, APOA1, APOF), hormonal status (e.g. IGFBP1, IGFBP2, SHBG, FGF21), or thermogenesis (e.g. LDLR, GHR, SHBG, CKB, LAMP2). A total of 129 proteins were associated with GRS_uncoupling_, but not with GRS_BFP_ (**Extended Data** Fig. 6, **Supplementary Table 13**), including neuropeptides (AGRP, NPY, BDNF), hormones (GCG, ADIPOQ), lipoprotein lipase (LPL), and myostatin (MSTN).

**Fig. 7:**
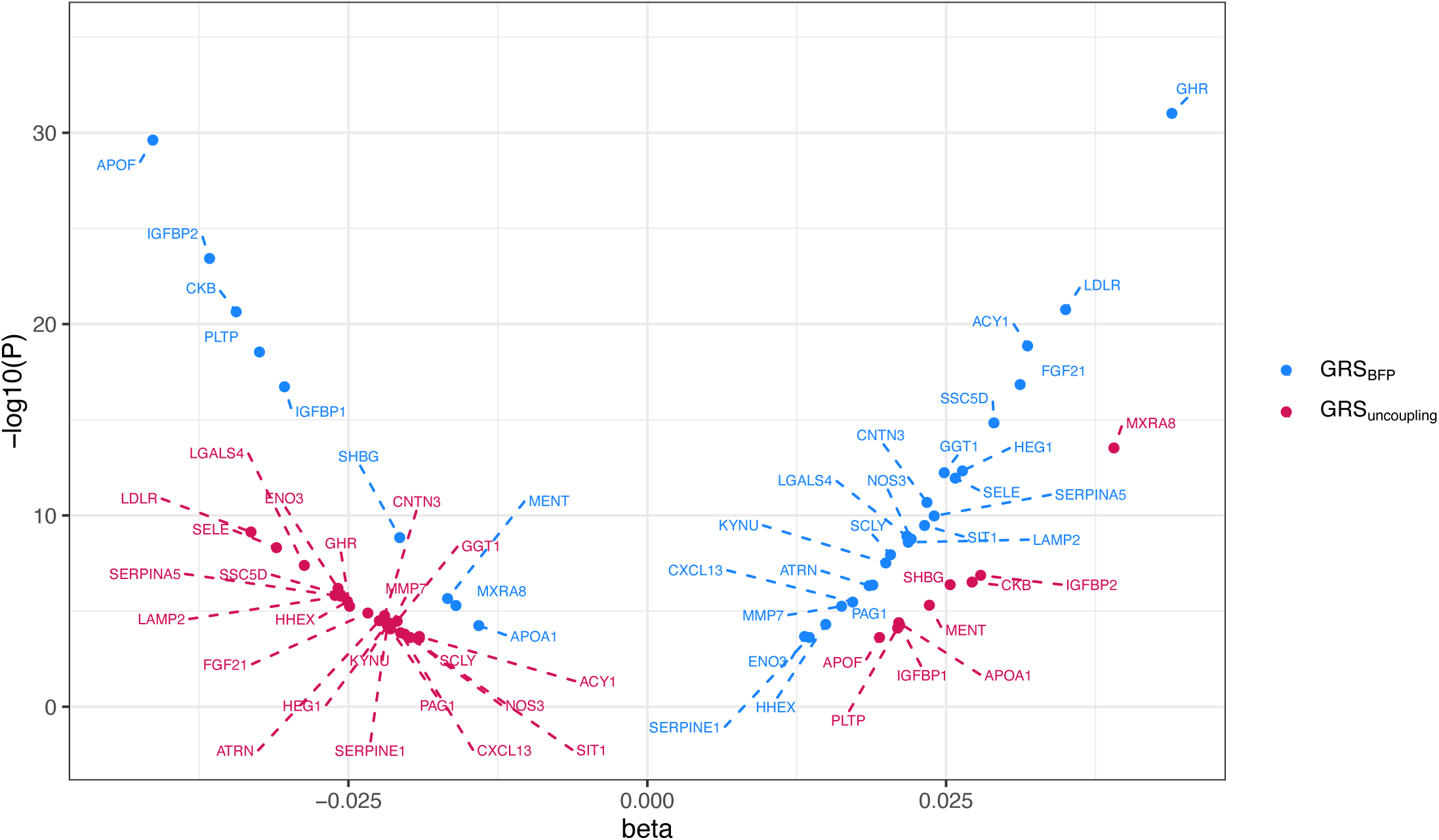
Plasma proteins (n=32) with directionally opposing associations with the body fat percentage- and uncoupling genetic risk scores in the UK Biobank. Estimated per 10 allele change effect sizes of GRS-protein associations in UK Biobank European ancestry population for GRS_uncoupling_ (in magenta) and GRS_BFP_ (in blue), for rank-based inverse-normal transformed Olink-derived plasma protein concentrations.

### Gene prioritization identifies genes implicated in various pathways

To identify putative causal genes within the 205 uncoupling loci, we used 14 bioinformatic and functional genomics tools. We prioritized the gene(s) most likely to be causal in each locus and ranked them based on the number of genomic tools that provided support for the given gene. Of the 1,623 candidate genes, 82 were prioritized by seven or more genomic tools and were considered high-scoring genes **(Methods, Supplementary Table 15).** The high-scoring predicted effector genes include genes previously described to be associated with opposite effects on obesity and cardiometabolic traits, such as *PPARG*^7,17–19^, *FAM13A*^9,11^, *PEPD*^11,19–21^, and *IRS1*^11,19,22,23^. The two highest scoring genes, prioritized by 11 tools, were *PCSK1* and *SMG6* **(Supplementary Table 15)***. PSCK1* encodes an enzyme (PC1/3) that cleaves prohormones, such as proopiomelanocortin, and its role in obesity has been well-established^24,25^. There is no obvious functional role for *SMG6* in cardiometabolic disease, and for neither *PCSK1* nor *SMG6* for their involvement in uncoupling of obesity from cardiometabolic health.

Other high-scoring genes have been implicated in adipose tissue expandability (*PPARG*^26,27^*, IRS1*^28–30^, *RSPO3*^31^*, FAM13A*^32^*, CTSS*^33,34^*, TIMP4*^35,36^*, PEPD*^19–21^*, JMJD1C*^37^*, CSK*^38,39^*, HLX*^40^*, MED19*^41^*, SENP2*^42^*, MLXIPL*^43,44^*, ARNT*^45,46^*, PIK3R1*^47^, PNPLA2^48^, insulin secretion and beta cell function (*PIK3R1*^49^*, GPRC5B*^50^*, MEF2D*^51^*, FBN1*^52^*, LDB1*^53^*, SENP2*^54^*, MAPT*^55^*, PBX1*^56^, beiging of white adipose tissue and brown adipose tissue function (*CSK*^39^*, SLC22A3*^57^*, SENP2*^58^*, MED19*^41^*, LDB1*^59^*, HLX*^40^*, CRHR1*^60^), and inflammation and fibrosis *(PEPD*^20,21^*, BCN2*^61^*, MST1*^62–64^*, GPRC5B*^65^*, MAFF*^66^*, CTSS*^34,67^*, NPEPPS*^68^*, CSK*^38^*, FBN1*^52,69^).

Many of these genes and pathways have been described before in the context of uncoupling^11^, but we also identify genes involved in pathways that have not previously been implicated, such as hepatic control of glucose homeostasis (*ARNT*^70^*, CTSS*^34,71^*, YWHAB*^72^*, FBN1*^73^ *, LDB1*^74^*, SENP2*^75^), hepatic lipid accumulation (*JMJD1C*^76^*, NPEPPS*^77^*, MLXIPL*^44,78^) and skeletal muscle growth and function (*PPP3R1*^79^, *CTSS*^80,81^*, CXXC5*^82^*, NPEPPS*^83^*, SENP2*^84^*, FBN1*^69^) **(Supplementary Table 16).**

Prioritized genes in loci that belong to the same cluster tend to share related pathways. For example, prioritized genes in cluster 7 are implicated in regional adipose expandability, including *FAM13A*^32^ and *RSPO3*^31^, consistent with lower WHR and improved lipid profile, being the defining characteristics for cluster 7. Another example is for cluster 3, characterized by lower systolic and diastolic blood pressure, which contains several genes implicated in beiging of white adipose tissue and brown adipose tissue function, including *CSK*^39^, *HLX*^40^, *LDB1*^59^, *MED19*^41^, and *SENP2*^58^. Brown adipose tissue has been previously linked to lower odds of cardiometabolic diseases including hypertension^85^. The overall protective cluster 8 has a more diverse biological basis with multiple genes implicated in adipose expandability, including *PPARG*^26,86^, *IRS1*^23,28–30,87^, *TIMP4*^36^, *CTSS*^34^, *ARNT*^45,46^, and *PIK3R1*^47^. *TIMP4* is also implicated in nutrient uptake^35^, and ARNT and CTSS in hepatic glucose control^70,71^. Therefore, our prioritized genes are implicated in known and/or novel pathways that plausibly contribute to the uncoupling of adiposity from cardiometabolic risk.

## DISCUSSION

Obesity is a highly heterogeneous disease that cannot be captured by one single adiposity trait. Here, we performed a multi-trait gene-discovery analysis to account for heterogeneity in cardiometabolic comorbidities. We designed uncoupling phenotypes that are continuous and range from high adiposity with a healthy cardiometabolic profile to low adiposity with an unhealthy cardiometabolic profile. GWASs of these new phenotypes identified 266 independent variants across 205 genomic loci where the adiposity-increasing allele is also associated with a lower cardiometabolic trait. A genetic score (GRS_uncoupling_) that aggregates the uncoupling effects of the 266 variants confirms the uncoupling of obesity from its cardiometabolic comorbidities, but also shows association with a higher risk of many other disorders, such as arthrosis, COPD, and asthma. Furthermore, the 266 variants cluster into eight groups, each representing a genetic subtype with a distinct cardiometabolic risk profile, pointing to specific, often peripheral, underlying pathways.

The genetic uncoupling score (GRS_uncoupling_) was associated with a healthier cardiometabolic profile, distinctly different from that of the genetic adiposity score (GRS_BFP_), which was associated with an increased cardiometabolic risk. The protective effects of the uncoupling score may be partially mediated through an association with a more favorable fat distribution characterized by a lower WHR and lower MRI-derived VAT/ASAT, in particular among women, compared to the adiposity score. These findings corroborate previous observations with greater power, including the sex-specific observations^9,10,12,88,89^. With such distinct cardiometabolic risk profiles, these genetic scores may help with an early risk stratification of individuals with obesity, subsetting those at risk of cardiometabolic comorbidities, allowing for a timely and personalized prevention, from those without such risk. This genetic risk stratification was already apparent in childhood and adolescence. Moreover, the uncoupling score was also associated with lower risk of prevalent and incident T2D and CHD in adulthood. However, while the cardiometabolic risk is reduced among individuals with a high uncoupling score, risk for diseases, such as cellulitis, arthrosis, sleep disorders, and phlebitis among others, is comparable to those with a high adiposity score, consistent with a previous report^90^. These observations underscore that the weight-bearing impact of a high body weight on overall health remains, even when cardiometabolic risk is lower.

The 266 lead variants for the uncoupling loci cluster into eight distinct subsets, based on their association profile with the adiposity and cardiometabolic traits, each representing a genetic subtype of obesity. In previous studies, we and others have identified three clusters of uncoupling loci^11,14^. Here, with many new uncoupling loci, our cluster analysis replicates the previous reported clusters with greater delineation and identifies several new clusters. For example, in previous studies, one of the uncoupling clusters identified was characterized by mediation through favorable fat distribution. We identified two such clusters (4 and 7), that, however, differ in the strength of associations with each of the phenotypes. Most of the new clusters are characterized by the fact that the adiposity increasing loci are associated with a specific subset of cardiometabolic traits. For example, the adiposity increasing loci in cluster 5 with a healthier glycemic profile, those in cluster 3 with lower blood pressure, and those in clusters 1, 6 and 8 are associated with lipid levels with three distinct profiles. Our findings extend current knowledge by underscoring that there is substantial heterogeneity among uncoupling loci. The clusters of loci represent distinct genetic subtypes that suggest a range of mechanisms underlying the uncoupling of obesity from its cardiometabolic comorbidities.

To gain further insight in the uncoupling mechanisms, we examined association of the genetic scores with plasma protein levels. We identified shared and distinct proteomic association signatures for the uncoupling score versus the adiposity score. For the vast majority (85%) of the 208 proteins associated with both genetic scores, the direction of the association was consistent across both scores. This suggests that, for these proteins, levels are driven by adiposity. For example, levels of leptin and adipsin/complement factor D, two adipokines known to be elevated in individuals with obesity irrespective of their cardiometabolic health status^91^, increased with the increase in both uncoupling and adiposity scores. A subset of proteins (15%) showed directionally opposite associations between the two scores, capturing health-driven effects. For example, higher plasma levels of IGFBP1 and IGFBP2 were associated with a higher uncoupling score; i.e. higher adiposity and improved cardiometabolic health, but with a lower adiposity score, representing lower adiposity and improved cardiometabolic health, corroborating previous reports demonstrating that lower levels of IGFBP1 and IGFBP2 are associated with hypertriglyceridemia and insulin resistance^92–94^. Also lower LDLR levels and higher SHBG levels were associated with a higher uncoupling score, consistent with a metabolically healthy state observed by others^95–100^. Several proteins were exclusively associated with a higher uncoupling score, such as ADIPOQ and LPL. ADIPOQ has been shown to be higher in metabolically healthy individuals, promoting insulin sensitivity and having cardioprotective and anti-inflammatory effects^91,101,102^, whereas LPL plays a role in triglyceride clearance and lipid distribution, potentially contributing to a metabolically healthy state^103,104^. Myostatin levels decreased with an increasing uncoupling score. Myostatin is considered a drug target for sarcopenia and muscle mass preservation in combination with weight-loss drugs^105,106^, implicating skeletal muscle mass and function in metabolic health^91^.

Follow-up analyses of the uncoupling loci highlight the importance of peripheral biology in uncoupling, in contrast to a role of the central nervous system in overall obesity. Tissue enrichment analysis for genes in uncoupling loci pointed to adipocytes and adipose tissue, cardiovascular, digestive, endocrine, and musculoskeletal systems, implicating pathways previously reported for uncoupling (insulin signaling, glucose homeostasis, lipid metabolism, immune and inflammatory response, adipose tissue biology)^7,11,15^, but also revealing new ones (e.g. vascular development, skeletal muscle development, liver development, kidney development, cartilage development, circadian rhythm, sex differentiation, and response to hypoxia). Even though the enrichment analyses were based on far fewer uncoupling loci, compared to adiposity loci, they reached much greater significance level for the uncoupling loci. This may be indicative greater heterogeneity among adiposity loci and/or greater homogeneity among the uncoupling loci.

To pinpoint candidate causal genes within each uncoupling locus, we established a bioinformatics and functional genomics gene prioritization pipeline. We identified genes previously implicated in uncoupling of adiposity and cardiometabolic risk^7,9,11,15,17–22^ and identified additional ones. The highest scoring genes fall into pathways that have been implicated before, thereby providing further support for biological processes such as adipose tissue expandability, fat distribution, insulin secretion and beta cell function, beiging of white adipose tissue and brown adipose tissue function. Other newly identified genes highlight emerging mechanisms, such as inflammation and fibrosis, hepatic glucose control and lipid accumulation, and muscle function. For example, liver specific ablation of *Jmjd1c*, a gene prioritized for rs10761785, which is associated with higher BMI, WHR, and lower LDL-C and total cholesterol levels, decreases lipogenesis and protects from insulin resistance despite obesity in mice^76^. Knockout of *Arnt*, a gene prioritized for rs10888393 and associated with increased BFP, high HDL-C, and low HbA1c, may have a tissue-specific role affecting adiposity and cardiometabolic health. Fat-specific *Arnt* knockout mice are leaner and protected against diet-induced glucose intolerance and obesity, whereas hepatocyte-specific *Arnt* knockout mice have increased fasting glucose and impaired glucose tolerance^45,46,70^.

Taken together, by designing continuous uncoupling traits, we substantially increased statistical power for discovery, resulting in a more than two-fold increase in the number of uncoupling loci identified. Gene prioritization efforts and pathway and protein analyses underscore the importance of a range of peripheral pathways in uncoupling. We provide further support for fat tissue expandability, insulin secretion and beta-cell function, beiging of white adipose tissue, inflammation and fibrosis, and also highlight mechanisms not previously implicated, such as hepatic lipid accumulation, hepatic control of glucose homeostasis, and skeletal muscle growth and function. Individuals with a high genetic uncoupling score, which aggregates the effects of the 266 lead variants, display a protective cardiometabolic risk profile despite having a higher risk of obesity. Their profile is distinct from that of individuals with a high overall adiposity score who have an increased cardiometabolic risk, and this risk stratification is already apparent in childhood and adolescence. The overall genetic uncoupling score and its eight derived sub-scores contribute to the genetic subtyping of obesity. These genetic subtypes may form the basis of subtype-stratified treatment, prevention and prognosis and may ultimately contribute to precision medicine in obesity.

## METHODS

### Study populations UK Biobank

The UK Biobank is a prospective cohort study with extensive genetic and phenotypic data, collected in approximately 500,000 individuals, aged between 40–69 years. Participants were enrolled from April 2007 to July 2010 at one of 21 assessment centers across the UK. Baseline information, physical measures, and biological samples were collected according to standardized procedures^107–109^. Questionnaires were used to collect health and lifestyle data. Study design, protocols, sample handling and quality control have been described in detail elsewhere^107–109^.

#### Ancestry

We restricted analyses to individuals of European ancestry, defined by using k-means clustering^110^. Briefly, we calculated principal components and their loadings for 488,377 genotyped participants based on the intersection of ∼121,000 QC’d variants with the 1000 genomes reference panel (phase 3 v5). We projected the 1000 genomes reference panel dataset on the PCA loadings from the UK Biobank. We then applied k-means clustering to the UK Biobank PCA and the projected 1000G reference panel dataset, pre-specifying 4 clusters. Individuals that clustered with the EUR 1000G cluster were assigned European ancestry.

#### Phenotypes

We analyzed 11 single traits; three adiposity traits: Body mass index (BMI), body fat percentage (BFP) and waist to hip ratio (WHR); and 8 cardiometabolic traits: total cholesterol (TC), LDL cholesterol (LDL-C), HDL cholesterol (HDL-C), triglycerides (TG), glucose, HbA1c levels, systolic blood pressure (SBP), and diastolic blood pressure (DBP). All phenotypic data used for analyses were collected at the baseline visit. BMI was calculated as weight (kg) divided by height squared (m2). WHR was created by dividing the waist circumference by the hip circumference. Individuals with waist and hip measurements <50 cm and >150 cm were removed. For individuals on lipids lowering medication, LDL-C was adjusted by dividing the LDL-C value by 0.7 and TC by dividing by 0.8. TC and TG were log transformed. For the GWAS analysis of glucose and HbA1c, individuals receiving diabetes medication, (N=4,697) and those with glucose > 15 mmol/l or HbA1c > 100 were excluded (N=804). SBP and DBP were created by calculating the average of two measurements at the baseline visit and adjusted for medication use by adding 15mm Hg on the SBP value and 10mm Hg on the DBP value. Upon exploring the data, we identified a subgroup of women recruited in one center, Sheffield, that deviated from the rest of the data. As a result, we excluded 121 women recruited at Sheffield and that had BFP > 55% and BMI > 40 kg/m2. Additionally, women who were pregnant at the time of recruitment were excluded. Finally, 452,768 participants (207,204 men and 245,564 women) were included in the analyses of lipid traits and 448,071 (204,377 men and 243,694 women) for glycemic traits.

#### Genotypes

Participants were genotyped on two arrays. The majority (N∼450,000) were genotyped using the UK Biobank Axiom Array and the remaining participants (N ∼50,000) were genotyped using the UK BiLEVE Array, which has >95% of the variants in common with the UK Biobank Axiom Array. Quality control, performed by the UK Biobank team, included testing for batch, plate, and array effects, Hardy-Weinberg equilibrium, and discordance across control replicates. Samples of poor-quality, with high missingness rate and heterozygosity, were removed^109^. Missing SNPs were imputed using the UK10K reference panel by the UK Biobank team.

### Atherosclerosis Risk in Communities (ARIC) Study

The ARIC study is a prospective cohort study of 15,792 individuals, including 11,478 whites and 4,314 African Americans, from four US communities (Forsyth County, NC; Jackson, MS; suburbs of Minneapolis, MN; and Washington County, MD). Participants aged between 45 to 64 years at baseline, recruited between 1987–1989, received extensive examinations, including medical, social, and demographic data. A detailed description of the ARIC study design was published elsewhere^111^. Adiposity and cardiometabolic traits considered in this study were measured at the baseline visit.

Incident CHD was ascertained through a combination of death certificate reviews, hospital records, and annual participant follow-ups to identify hospitalizations and deaths occurring during the prior year^112^. Incident CHD cases were defined as definite fatal CHD, definite or probable myocardial infarction (MI), silent MI between examinations as determined by ECG, or coronary revascularization. T2D cases were identified at baseline and during follow-up visits using glucose measurements, self-reported physician diagnosis of type 2 diabetes or use of diabetes medication. T2D was defined in accordance with World Health Organization (WHO) guidelines as a fasting serum glucose ≥7.0 mmol/L, a non-fasting serum glucose ≥11.0 mmol/L (when fasting samples were unavailable), or the use of blood glucose-lowering medications. Individuals with T2D at baseline were removed from this analysis.

Blood was drawn for DNA extraction at the baseline exam. Genotyping within ARIC was performed on the Affymetrix 6.0 DNA microarray (Affymetrix) and genotype data that passed quality control filters was imputed into the 1000 Genomes phase 3 reference data using IMPUTE version 2.3.2^113,114^. The ARIC study was approved by the institutional review boards at each site, and written informed consent was obtained from all study participants.

#### Bio*Me* Biobank

Bio*Me* is an ongoing electronic medical record-linked biobank with more than 60,000 patients enrolled through the Mount Sinai Health System in New York. Bio*Me* is a multiethnic biobank comprising individuals of African, Hispanic, European, Asian, and other ancestries. Genotyping data on the Global Screening Array (GSA-24v1-0_A1) is available for 32,595 individuals. The data was cleaned for duplicate samples, discordant sex, heterozygosity rate that exceeded 6 SD from the population mean, call rate<95% at the site and individual level, and deviation from Hardy Weinberg equilibrium. After QC, 31,705 individuals and 604,869 variants were retained. Imputation of the GSA array was performed using impute2^115^ using the 1000 Genomes reference panel. In BioMe Biobank, CHD cases were identified using ICD-9 and ICD-10 codes, procedure codes for bypass surgery or percutaneous transluminal coronary angioplasty (PTCA), or documentation of abnormal cardiac catheterization. T2D cases were defined using the eMERGE phenotyping algorithm^116^. Baseline was defined as first outpatient visit after 01/01/2011, with at least one year enrolled in the Mount Sinai Health system. Individuals with CHD or T2D cases occurred before or within one year of enrollment were classified as prevalence cases and excluded from analyses.

### HOLBAEK

The HOLBAEK study consists of two Danish cohorts: a cohort from the Children’s Obesity Clinic of the Holbaek Hospital, comprising children and adolescents with a BMI at or above the 90th percentile (BMI standard deviation score (SDS) ≥ 1.28, i.e., overweight or obesity) based on Danish reference standards^117^; and a population-based cohort recruited from schools in 11 municipalities across Region Zealand^118^. In the obesity clinic cohort, anthropometrics were measured at clinical examinations, whereas the population-based group was assessed in a mobile laboratory by medical professionals. Details on the cohort and phenotypic data are published elsewhere^119^. We considered 23 traits for our cross-sectional analyses (5 binary, 18 continuous). BMI SDS was derived using least mean squares method, referenced against Danish reference standard^117^. Waist-to-height ratio (WtHR) SDS, and waist-to-hip ratio (WHR) SDS were calculated based on age- and sex-specific reference values from NHANES^120^. Obesity was defined as having a BMI SDS ≥ 2.33 (i.e., 99th percentile and above^121^). Hyperglycemia, insulin resistance, dyslipidemia, and hypertension were defined according to published guidelines^122–125^. HOMA-IR was calculated as (insulin mU l^−1^ × glucose mM)/22.5. Exclusion criteria for the current analyses included individuals younger than 5 or older than 19 years of age, those taking medications for obesity or diabetes, participants meeting T2D criteria based on fasting plasma glucose levels ≥7.0 mmol/L or HbA1c ≥48 mmol/mol, and individuals without genotyping data.

The final number of participants of European ancestry analyzed was 1,646 for the obesity clinic cohort (45% boys, median age 11.7y [Q1 9.6y; Q3 14.0y]) and 1,811 for the population-based cohort (43% boys, median age 11.6y [Q1 8.9y; Q3 14.5y])

#### Genotype data

Genotyping in the HOLBAEK study was conducted using Illumina Infinium HumanCoreExome-12 v1.0 or HumanCoreExome-24 v1.1 BeadChips, analyzed on the Illumina HiScan system. Genotype calling was performed using the Genotyping Module (v1.9.4) within GenomeStudio software (v2011.1; Illumina). Phasing was done with EAGLE2 (v2.0.5), and imputation was carried out using PBWT to the Haplotype Reference Consortium (HRC1.1) via the SANGER imputation server. As HRC1.1 does not include insertions and deletions and does not fully overlap with imputed UK Biobank (UKB) genotype data, up to 20% of GRS_BFP_ and GRS_uncoupling_ variants were not available. We therefore identified high-LD (R2>0.8) proxies, based on LD information from 20,000 unrelated (KING<0.0884), randomly sampled UKB participants. These participants self-identified as ’White British’ and clustered with this group in principal component analysis (PCA). Genotype QC for this reference panel included filtering on SNP missingness <5% and INFO>0.3, and we excluded participants with sex chromosome anomalies, sex discrepancies, heterozygosity outliers, and genotype call rate outliers. Proxies were identified for 123/142 missing variants for GRS_BFP_ and 56/58 missing variants for GRS_uncoupling_, with a median R^2^_UKB_ of 0.99. The final GRSs therefore included 628 and 264 variants, respectively, with a median INFO_HOLBAEK_ of 0.98. The GRSs were scaled to per 10 allele change.

#### Statistical analysis

Using linear and logistic regression, we assessed the association between both GRSs and the 23 outcome traits, adjusting for age, sex, four PCs, and genetic batch (n=3). The continuous traits were log transformed (except for BMI-, WHR-, WHtR-, SBP-, and DBP-SDS) and then standardized to unit variance and standard deviation of one. All analyses were stratified by cohort (obesity clinic/population) and estimates pooled using inverse-variance weighting. In addition, all analyses were further stratified by sex (**Supplementary Table 11**). P-values were adjusted using Benjamini-Hochberg correction across the 23 traits.

### GWAS analyses

Our GWAS analyses aimed to identify variants that uncouple adiposity from its comorbidities. As such, we used the 11 single traits: three adiposity traits: BMI, BFP, and WHR; and 8 cardiometabolic traits: TC, LDL,-C HDL-C, TG, glucose, HbA1c, SBP, and DBP. First, we derived residuals for each of the single traits, for men and women separately, using linear regression analyses adjusting for age, age^2^, genotyping array, and sequencing center. Next, the distributions of residuals of the 11 single traits were inverse normalized. The derived SD-scores have a mean of 0 and a SD of 1. We then created pairwise composite traits with one of the anthropometric traits and one of the cardiometabolic traits by subtracting the SD-scores of the cardiometabolic trait (TC, LDL-C, HDL-C, TG, glucose, HbA1c, SBP and DBP) from those of the adiposity traits (BMI, BFP, and WHR), resulting in 24 bi-traits. For example, a BMI-TC bi-trait is created as follows: BMI_sdcores_ - TC_sdcores_ **(Extended Data** Fig. 1). For any given individual, a positive score for the BMI-TC bi-trait indicates that the person has a relatively higher BMI compared to their TC levels, whereas a negative score means that they have a relatively lower BMI compared to their TC levels. For BMI as the adiposity measure, we have 8 bi-traits: BMI-TC, BMI-LDL-C, BMI-HDL-C, BMI-TG, BMI-glucose, BMI-HbA1c, BMI-SBP, BMI-DBP. Similarly, for BFP: BFP-TC, BFP-LDL-C, BFP-HDL-C, BFP-TG, BFP-glucose, BFP-HbA1c, BFP-SBP, BFP-DBP, and for WHR: WHR-TC, WHR-LDL-C, WHR-HDL-C, WHR-TG, WHR-glucose, WHR-HbA1c, WHR-SBP, WHR-DBP.

As such, we performed 70 mixed model GWAS tests analyzing 24 bi-traits and 11 single traits within men and women separately (35 traits x 2) using BOLT-LMM^126^ adjusting for the first 10 principal components.

Variants with MAF<0.1% were removed from the analyses. For imputed variants, an INFO score threshold=0.3 was used. For each trait, we used METAL to meta-analyze the GWAS results of men and women using fixed effects^127^. Using LD score regression (LDSC), we observed mild inflation, with LDSC intercepts ranging between 1.09 and 1.18 and LDSC ratios indicating 10-13% of the inflation observed can be ascribed to causes other than a polygenic signal. Therefore, we applied genomic control by adjusting the standard error for the LDSC intercept. Specifically, for each trait, we used the corrected standard error: SE_corrected_= SE_GWAS_ x sqrt (LDSC intercept) of the sex-specific GWAS as the SE column in the inverse-variance weighted meta-analysis of men and women. The new corrected LDSC intercepts ranged from 1.03 to 1.08. LDSC ratio indicated that no more than 5.7% of the inflation observed can be ascribed to causes other than a polygenic signal.

#### Conditional analyses

To identify additional independent signals in associated loci for the 24 bi-traits, we used GCTA^128^. We performed approximate joint and conditional SNP association analyses in each locus, which takes into account LD between SNPs. For each locus, we defined a 2 Mb region encompassing 1 Mb on both sides of the lead SNP. Lead SNPs (P < 5×10^-10^) identified in known long-range high-LD regions were treated as a single large locus in the GCTA analysis^129^. We used unrelated European ancestry participants from the UK Biobank as the reference sample to acquire conditional P-values for association. Conditional independent variants that reached P < 5×10^-10^ were considered as index SNPs. We additionally restricted to SNPs that were genome-wide significant (P < 5×10^-10^) in the original summary statistics.

#### Identification of genome-wide significant loci

Our genome-wide significance threshold of P<5×10^-10^ accounted for the analyses of 24 bi-traits, across women and men. To identify genome-wide associated loci and their respective lead SNPs, we proceeded as follows. We started with the independent variants that resulted from the conditional analyses. To define a locus associated with increased adiposity and protective effects on cardiometabolic traits, we retained only variants for which the single-trait GWASs for both traits reached marginal significance, defined as P<10^-4^, and for which the association was opposite to the established phenotypic correlation. For example, for BMI and LDL-C, we selected variants for which the BMI-LDL-C bi-trait reached genome-wide significance, and subsequently extracted the variants for which the single-trait associations with BMI and with LDL-C reached P<10-4 and their direction of association was opposite of the established phenotypical positive association. As such, we identified 1,103 association signals across the 24 bi-traits. Next, we applied a Bayesian divisive clustering algorithm, HyPrColoc^130^ to determine, for each of the 1,103 association signals, whether the associations across the bi-traits and both single traits co-localize. In the above example, we would only keep loci of which the associations co-localize across the BMI-LDL-C bi-trait, BMI, and LDL-C. HyPrColoc is a deterministic Bayesian algorithm that, for a given genomic region, identifies clusters of traits which colocalize at distinct causal variants^130^. The algorithm also allows for sample overlap for the tested traits and corrects for it. As such, we provided the following input files to HyPrColoc upon testing the co-localization for each of the 1,103 associations: [1] a file with the beta values of all the variants to be tested and the traits in consideration, [2] another file with their SEs; [3] a 3x3 matrix denoting the phenotypic correlation between the bi-trait and its corresponding pair of single traits estimated from the UK Biobank data; [4] an LD matrix for all variants within 1mb region around the lead SNPs; and [5] a 3x3 matrix with all values equal to 1. We used a 1Mb region around the input variant (0.5Mb on each side). Therefore, of 1,103 association signals, we found that 602 association signals corresponding to 425 unique SNPs co-localized across the bi-traits and their corresponding single traits. We subsequently inspected LocusZoom plots for potential overlap between independent loci across the traits and for missed co-localized association signals because HyPrColoc assumes only one “causal” variant per region. This manual inspection led to either narrowing or widening the genomic region of 32 co-localized association signals and performing HyPrColoc again to identify other co-localized variants that were missed.

If more than one variant in the same region was retained for the different traits, we chose the lead variant based on the following criteria. If both variants were in high LD (r2>0.9), we randomly picked one of the two. Otherwise, if both variants were significantly associated with the same or similar traits, then we chose the variant that had lower P-values for a larger number of traits. However, if two variants in the same region were each associated with a bi-trait that represents different categories of cardiometabolic traits, we kept both variants even though they were in the same locus. For example, if one variant was associated with BMI-SBP and another with BMI-HbA1c, then we kept both, as the two may be contributing to the uncoupling of adiposity and co-morbidities via different mechanisms. We finally retained 266 unique variants in 205 genomic loci.

### Cluster analysis

We used the Noise-Augmented von Mises-Fisher Mixture model (NAvMix) algorithm to cluster the 266 lead variants based on their association with the single traits^131^. Briefly, noise-augmented directional clustering clusters variants based on their proportional associations with different traits. The algorithm outputs a probability of membership for each datapoint (i.e. variant) to belong to each cluster. Each datapoint is then assigned to the cluster for which it has the highest probability. The procedure is repeated for a varying number of clusters and the final number of clusters is chosen based on the Bayesian Information Criterion (BIC). NAvMix outputs a noise cluster that includes data points (i.e. variants) that do not belong to any cluster and are thus considered outliers. We used the effect size (beta) of the association of each of the 266 lead variants with each of the eleven single traits as input. We used BFP as reference, assuming a positive direction of effect to facilitate comparison across traits. The associations with other traits were expressed using the BFP-increasing allele as the effect allele and the BFP-decreasing allele as the alternate allele.

### Genetic risk scores (GRSs) and their association with anthropometric and cardiometabolic traits, and phenome-wide association study

We constructed GRSs for all 266 identified variants combined (GRS_uncoupling_) and for each of the eight clusters separately (GRS_1_-GRS_8_) in 373,747 unrelated individuals of European ancestry from the UK Biobank. We also generated a GRS for body fat percentage (GRS_BFP_) based on 647 lead variants that reached P < 5×10^-9^ (clumped for r^2^<0.1 in 1mb region, MHC region removed) in our single trait GWAS for BFP in the UK Biobank. All of the 10 GRSs were weighted by the effect size estimated for BFP from the GWAS that we performed in the current study. The GRSs were rescaled to per 10 allele change^132^ to enable comparison across GRSs that consist of different number of SNPs. Cluster 2 was identified by the clustering analysis to be associated with lower WHR. For all analyses that followed the clustering analyses, the effect and reference alleles of genetic variants that were used to construct GRS_2_ were flipped in order to reflect a profile of higher adiposity and facilitate comparison with the other clusters.

To test the GRSs’ associations with anthropometric and cardiometabolic traits, we performed linear regression analyses for 21 traits, including height, WHR, hip circumference, waist circumference, BFP, fat-free mass index (FFMI, computed as whole body fat-free mass divided by height squared), BMI, gynoid fat percentage, android fat percentage, MRI measured visceral adipose tissue volume (VAT), abdominal subcutaneous adipose tissue volume (ASAT),, VAT/ASAT ratio, liver proton density fat fraction (liver fat), SBP, DBP, HDL-C, TG, LDL-C, TC, glucose and HbA1c. Each of the traits was adjusted for age, sex, genotype array and study site and the first ten principal components in a linear regression model. The resulting residuals were transformed to approximate normality using inverse normal rank scores before the association testing.

In addition, we performed phenome-wide association (PheWAS) analyses in unrelated individuals of European ancestry from the UK Biobank using the PHEnome Scan ANalysis Tool (PHESANT)^133^. Analyses were performed using a linear or logistic regression for continuous and binary outcomes, respectively, using the following covariates: age at enrollment, sex, genotyping array, and the first 10 genetic principal components (PCs).

### Statistical analysis in ARIC and Bio*Me*

We generated 10 GRSs: GRS_uncoupling_,GRS_1_-GRS_8_, and GRS_BFP_ in 9,240 unrelated European ancestry participants of the ARIC study. For the continuous traits, we created rank-based inverse normal transformed traits adjusting for age, sex, and the first ten PCs. We analyzed adiposity and cardiometabolic traits including hip circumference, waist circumference, WHR, BMI, SBP, DBP, HDL-C, TG, LDL-C, TC, and glucose in ARIC. We performed linear regression analyses to test the association of each GRS with the continuous traits. For analyses of incident T2D and CHD, we generated 10 GRSs: GRS_uncoupling_ and GRS_BFP_ in 23,208 unrelated individuals of European (N=8,985), Hispanic (N=7,984) and African (N=6,239) ancestry from the Bio*Me* Biobank. We tested the association of each GRS with incident T2D and CHD in Bio*Me* and ARIC using a cox proportional hazard model after adjusting for age, sex, ancestry (for Bio*Me*) and 10 principal components. Bio*Me* and ARIC association results were meta-analyzed using inverse variance weighted meta-analysis.

### Plasma proteomic characterization of the GRSs

We assessed the association of GRS_uncoupling_,GRS_1_-GRS_8_, and GRS_BFP_ with Olink-derived plasma protein measurements in the UK Biobank. This analysis included 30,271 unrelated individuals of European ancestry from the random baseline sample selected by the UKB Pharma Proteomics Project (UKB-PPP)^16^, specifically those included in Olink batches 1-6. After excluding three proteins with >25% missing data (PCOLCE (62%), NPM1 (73%), GLIPR1 (>99%)), we included 2,920 proteins with a median missingness of 2.8%. Measurements below the limit-of-detection (LOD) were included, in line with Olink’s recommendations [https://olink.com/faq/how-is-the-limit-of-detection-lod-estimated-and-handled/]. Using linear regression, we assessed the association between the 10 GRSs (scaled to a per 10 allele change) and protein measurements (rank-based inverse normal transformed). Covariates included age at measurement, age squared, sex, UK Biobank assessment center, 10 PCs, genotyping array, Olink batch, fasting time at measurement (hours), and time between measurement and processing of the sample by Olink (years). Benjamini & Hochberg adjustment was applied to control the false discovery rate at 0.005 (0.05/10*GRS) across the 29,200 protein∼GRS associations. To distinguish between adiposity- and health-driven associations, we grouped proteins that showed evidence of association with both GRS_BFP_ and one or more of the adiposity-uncoupled GRSs, based on their directional concordance. Moreover, we assessed which proteins uniquely associated with any of the adiposity-uncoupling GRSs, but not GRS_BFP_.

### Gene prioritization analysis

To identify the likely causal gene(s) within each of the identified loci, we used a combination of up to 14 bioinformatics and functional genomics methods. In addition to the annotated “nearest gene”, we used a “coding proxy” approach, five bioinformatics tools, and leveraged *in vitro* adipogenic differentiation dataset with seven measures. For each gene, we summed the number of methods for which it was prioritized. Genes with a score ≥ 7 were prioritized. The methods used towards the gene prioritization score are described below.

### Bioinformatics tools

#### Nearest gene

We used the nearest gene as predicted by Ensembl Variant Effect Predictor (VEP)^134^.

#### Coding proxy

For a given lead SNP, we considered variants in high linkage disequilibrium (LD) (r2>0.8) within a 1Mb window. If one or more of those variants was a coding variant, then the gene(s) in which those coding variants lie were prioritized. We annotated the variants in (VEP). A coding variant was defined as any variant with the following annotations: synonymous_variant, missense_variant, inframe_insertion, inframe_deletion, stop_gained, frameshift_variant, splice_donor_variant, and splice_acceptor_variant.

#### ABC-max

We used FUMA^135^ to select all SNPs in high LD (r2>0.8) using as reference panel UKB release 2b 10K Europeans. We intersected the total of 10,095 SNPs (lead SNPs and proxies in high LD) with enhancers and target genes predicted by the Activity-by-Contact (ABC) model^136^ in the following tissues: adipose, adrenal gland, astrocytes, pancreas, cardiac muscle cell, coronary artery, smooth muscle cell of coronary artery, heart ventricle, hepatocyte, liver, spleen, skeletal muscle myoblast and thyroid gland. Intersection of SNPs and enhancers was done using function *intersect* from bedtools v2.29.2^137^.

#### PoPS

We used Polygenic Priority Score v0.1^138^, a similarity-based algorithm that uses a broad range of omics data, including scRNA-seq. We used a reference panel of 10,000 randomly selected subjects from the UK Biobank and retrieved the gene with the highest PoPS score in each associated loci.

#### Data-driven Expression Prioritized Integration for Complex Traits (DEPICT)

We used summary statistics generated from the association analyses of the 266 lead SNPs with body fat percentage as input for DEPICT with default parameters^139^. DEPICT is an integrative tool that uses transcriptome expression (microarray data), pathways and protein-protein interactions to prioritize the most likely causal genes and highlight enriched tissues and pathways. **fastENLOC:** We colocalized associated loci with eQTLs from GTEx v8^140^ using fastENLOC^141^, prioritizing genes affected by eQTLs colocalizing at regional colocalization probability (RCP) >0.1. We colocalized lead SNPs and variants in high LD (r2>0.8) with eQTLs in adipose tissue (subcutaneous and visceral), pituitary, brain cortex, brain hypothalamus, brain hippocampus, brain amygdala, adrenal gland, thyroid gland, liver, kidney, pancreas, skeletal muscle, salivary gland and heart (atrial appendage and left ventricle).

### CS2G depot-specific gene prioritization

We intersected the 266 associated variants and their proxies (r2>0.8) with depot-specific chromatin accessible signals in subcutaneous and visceral human primary adipose-derived mesenchymal stem cells (AMSCs). We then performed combined S2G strategy CS2G^142^ to predict the effector genes. We reported the effector genes with a cutoff of 0.05 on the 1000 Genome and UK Biobank scores. The detailed protocol for AMSC proliferation, induction and differentiation is outlined in Laber *et al*.^143^. Overall, AMSCs were obtained from subcutaneous and visceral adipose tissue from patients undergoing a range of abdominal laparoscopic surgeries^143^ and isolated as previously described^144^. For a subset of donors, purity of AMCSs was assessed as previously described^145^. Each participant gave written informed consent before inclusion and the study was approved by the ethics committee of Technical University of Munich (Study No. 5716/13). Cells were introduced (counted as differentiation day 0) and put into differentiation for 14 days until fully differentiated. Samples are collected at differentiation day 14. Donor genotyping, SNP QC as well as the genotype imputation were performed as previously described^146^. Nuclei and library preparation for AMSC ATAC-seq were performed as previously described^146^. ATAC peaks were called by MACS3 (v3.0.0). After peak calling, narrow peaks from all the samples (Subcutaneous AMSCs n=15, visceral AMSCs, n=14) were first combined, then the overlapped intervals were merged into single interval using bedtools (version v2.30.0) (function bedtools merge -I)^137^.

### Overlap with eQTL data from GTeX

For the association and scoring of SNPs with genes we made use of eQTL data from GTEx (version 8) for subcutaneous and visceral adipose tissue as well as enhancer capture HiC data (“GSE140782_ECHiC.txt.gz” 10.1038/s41588-020-0709-z), DNase-seq based chromatin accessibility (“GSE113253_DNase_processed_data.tar.gz” 10.1038/s41588-019-0359-1), and gene expression data (“GSE113253_GeneExpr_BM.txt.gz” 10.1038/s41588-019-0359-1) of hBM-MSC-TERT4 cells subjected to adipocyte differentiation *in vitro*.

Specifically, the lead SNP or a proxy SNP, that was determined using the R package LDlinkR (10.3389/fgene.2020.00157) with a threshold of R^2^ > 0.8, had to fulfill at least one of the four criteria: 1) overlapping with an eQTL in visceral adipose tissue; 2)overlapping with an eQTL in subcutaneous adipose tissue; 3) overlapping with a genomic region that is linked by the enhancer capture data to a promoter region; or 4) overlapping with a DNase1 hypersensitive region. If only the latter case was true, the closest transcription start site was chosen to be the candidate gene. Overlap was determined using the R package GenomicRanges.

For each unique combination of lead SNP and putative candidate gene, we set up a score based on an overlap of the proxy SNPs with an eQTL, overlap of the proxy SNPs with a DNase1 hypersensitive site and its change in accessibility during adipocyte differentiation, overlap of the proxy SNPs with an enhancer region contacting the promoter of the candidate gene, and the expression of the candidate gene and its significant change during adipocyte differentiation.

### Tissue and Gene set enrichment

Tissue and gene set enrichment were performed by DEPICT, using summary statistics generated from the analysis on body fat percentage as input with default parameters^139^. Both were performed on all uncoupling loci and on cluster-specific loci. Only gene sets with at least 10 genes were included. We restricted our analyses to the Gene Ontology, KEGG, and REACTOME pathways terms. We used FDR<0.05 as a threshold for significance when considering all the 266 variants. For the cluster-specific analysis, since the clusters have a small number of variants and therefore less power, we used P<0.05 as a threshold from the “Nominal P value” output from DEPICT regardless of the FDR value.

## Data Availability

All data produced in the present work are contained in the manuscript.

## ACKNOWLEDGEMENTS

The Novo Nordisk Foundation Center for Basic Metabolic Research is an independent research center at the University of Copenhagen, partially funded by an unrestricted donation from the Novo Nordisk Foundation (NNF23SA0084103; NNF18CC0034900). This work is supported by the Novo Nordisk Foundation (NNF21SA0072102), UM1DK126185, R01DK102173. M.C. is supported by the Weissman Davis and Titlebaum MGH Research Scholar award. R.J.F.L. is supported by grants from the National Institutes of Health (R01DK107786; R01DK110113; R01HG010297; R01HL142302; R01DK075787; R01HL156991; U01HG011723; R01DK123019; R01HL151152; R01HL158884), the Danish National Research Fund (DNRF161) and the Novo Nordisk Foundation (NNF20OC0059313). V.D.O. is supported by a grant from the Danish Diabetes and Endocrine Academy, funded by the Novo Nordisk Foundation (NNF22SA0079901). J.C.H has received honoraria for expert roles from Novo Nordisk and Rhythm Pharmaceuticals. Dr Holm provides training and treatment of obesity.

The Atherosclerosis Risk in Communities study has been funded in whole or in part with Federal funds from the National Heart, Lung, and Blood Institute, National Institutes of Health, Department of Health and Human Services, under Contract nos. (75N92022D00001, 75N92022D00002, 75N92022D00003, 75N92022D00004, 75N92022D00005). The authors thank the staff and participants of the ARIC study for their important contributions. The datasets used for the analyses described in this manuscript were obtained from dbGaP under accession phs000223.

The Mount Sinai Bio*Me* Biobank is supported by The Andrea and Charles Bronfman Philanthropies and by Federal funds from the NIH (U01HG00638001; U01HG007417; X01HL134588). Furthermore, analyses were in part supported through the computational and data resources and staff expertise provided by Scientific Computing and Data at the Icahn School of Medicine at Mount Sinai and by the Clinical and Translational Science Awards (CTSA) grant UL1TR004419 from the National Center for Advancing Translational Sciences. Research reported in this publication was also supported by the Office of Research Infrastructure of the National Institutes of Health under award number S10OD026880 and S10OD030463. The content is solely the responsibility of the authors and does not necessarily represent the official views of the National Institutes of Health.

## Extended Data Figures

**Extended Data Fig. 1. An example illustrating how the new bi-traits are created.** Pairwise difference between BMI and TC z-scores results in a new normally distributed bi-trait BMI-TC.

**Extended Data Fig. 2. Sex-specific associations of genetic risk scores with anthropometric and cardiometabolic traits in the UK Biobank.** A. Estimated per 10 allele change effect sizes of GRS–trait associations in UK Biobank European ancestry population for GRS_uncoupling_ (men in light magenta, women in dark magenta) and GRS_BFP_ (men in light blue, women in dark blue). B-C. Estimated per 10 allele change effect sizes of GRS–trait associations in UK Biobank European ancestry population for each cluster specific GRS (GRS 1-8, men in light magenta, women in dark magenta). The dashed circles are labeled ’0’, indicating no association between each GRS and the trait. Points outside the circle represent positive GRS– trait associations, while those inside represent negative associations.

**Extended Data Fig 3. Associations of genetic risk scores with anthropometric and cardiometabolic traits in the HOLBAEK study.** A. Estimated per 10 allele change effect size of GRS-anthropometric trait associations for GRS_uncoupling_ (in magenta) and GRS_BFP_ (in blue). B-C. As A, but for associations with continuous and binary cardiometabolic traits, respectively. Continuous traits were standardized to mean 0 and SD 1. Dashed circles in A and B indicate Beta=0, and the dashed line in C indicates odds ratio=1. Points outside the dashed circles in A and B represent positive GRS–trait associations, while those inside represent negative associations. *analysis restricted to the population-based cohort only.

**Extended Data Fig. 4. Pathways enriched for GRS_uncoupling_ and GRS_BFP_ loci.** REACTOME, GO, and KEGG pathways with nominal P < 0.01 from DEPICT were grouped into broad pathway categories to enable visualization and plotted for each of GRS_uncoupling_ and GRS_BFP_. The width of each category is proportional to the number of pathways in that category. Redundant pathways were removed. Full results are presented in Supplementary Table 12.

**Extended Data Fig. 5: Plasma proteins (n=176) with directionally consistent associations with the body fat percentage- and uncoupling genetic risk scores in the UK Biobank.**

Estimated per 10 allele change effect sizes of GRS-protein associations in UK Biobank European ancestry population for GRS_uncoupling_ (in magenta) and GRS_BFP_ (in blue), for rank-based inverse-normal transformed Olink-derived plasma protein concentrations.

**Extended Data Fig. 6: Plasma proteins (n=129) which associate with the uncoupling-but not the body fat percentage genetic risk score in the UK Biobank.** Estimated per 10 allele change effect sizes of GRS-protein associations in UK Biobank European ancestry population for GRS_uncoupling_ (in magenta) and GRS_BFP_ (in blue), for rank-based inverse-normal transformed Olink-derived plasma protein concentrations.

