## Extended data for "Genetic subtyping of obesity reveals biological insights into the uncoupling of adiposity from its cardiometabolic comorbidities"

**Extended Data Fig. 1. An example illustrating how the new bi-traits are created.**

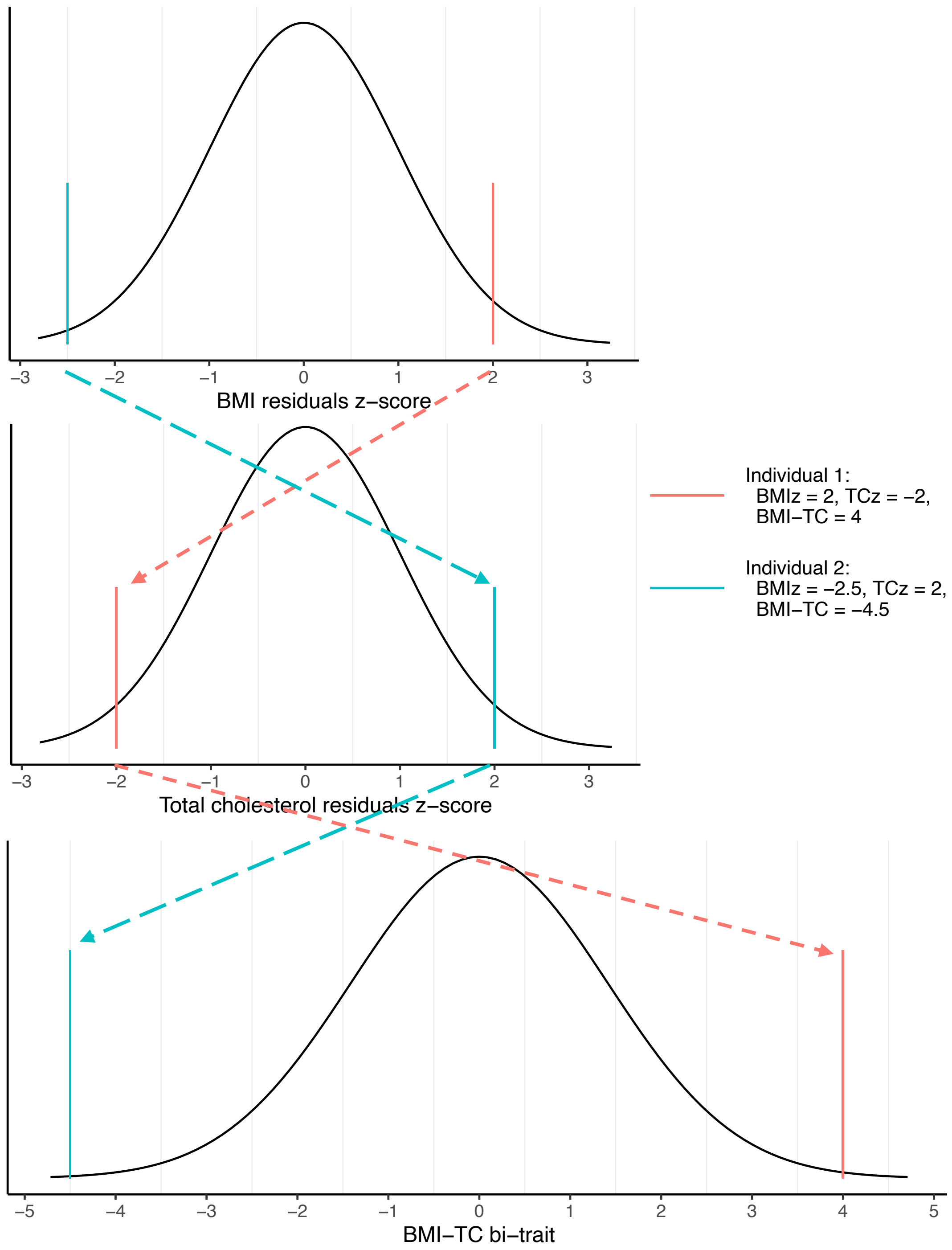

**Extended Data Fig. 2. Sex-specific associations of genetic risk scores with anthropometric and cardiometabolic traits in the UK Biobank.**

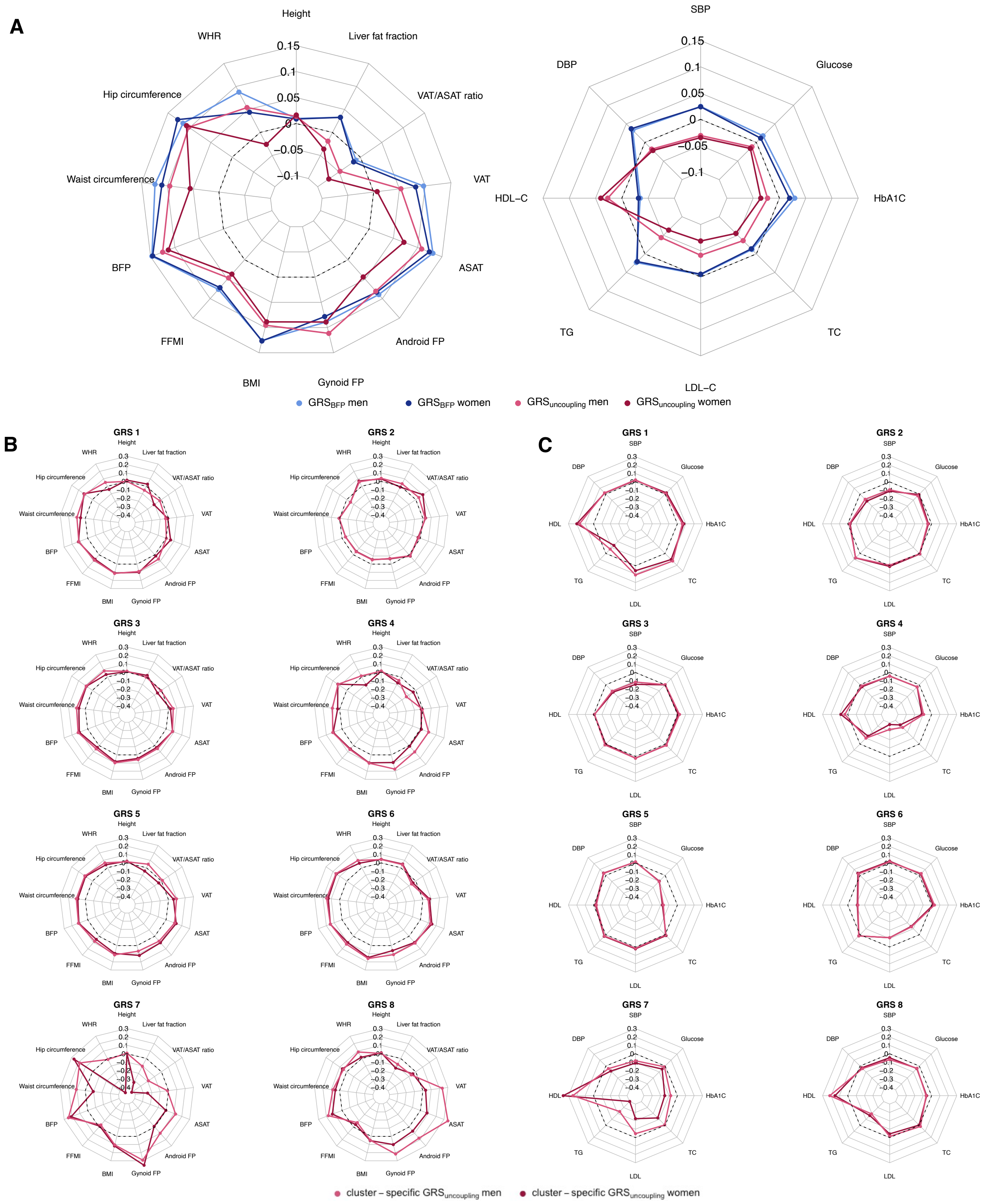

**Extended Data Fig. 3. Associations of genetic risk scores with anthropometric and cardiometabolic traits in the HOLBAEK study**

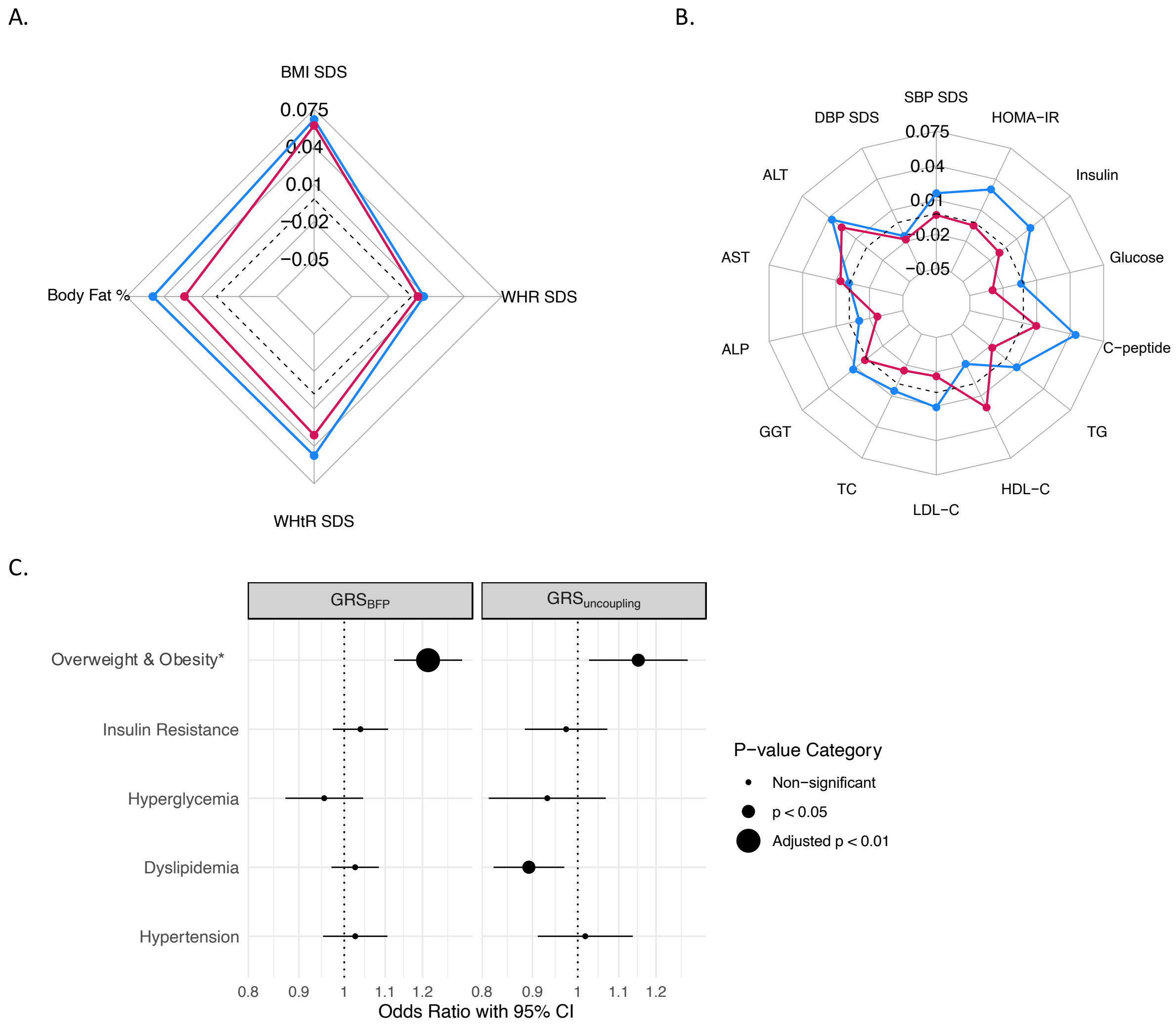

**Extended Data Fig. 4. Pathways enriched for $GRS_{uncoupling}$ and $GRS_{BFP}$ loci.**

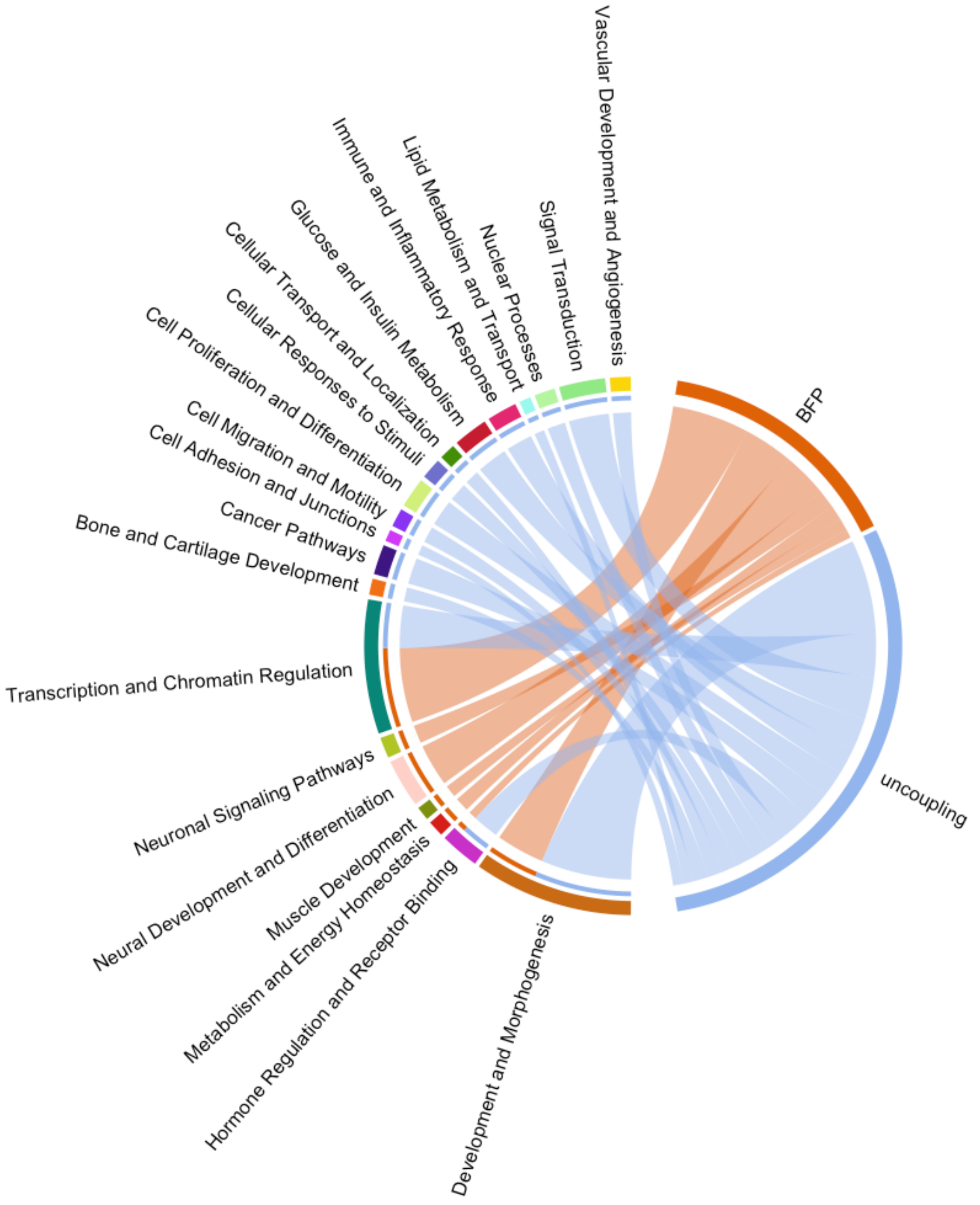

**Extended Data Fig. 5: Plasma proteins (n=176) with directionally consistent associations with the body fat percentage- and uncoupling genetic risk scores in the UK Biobank.**

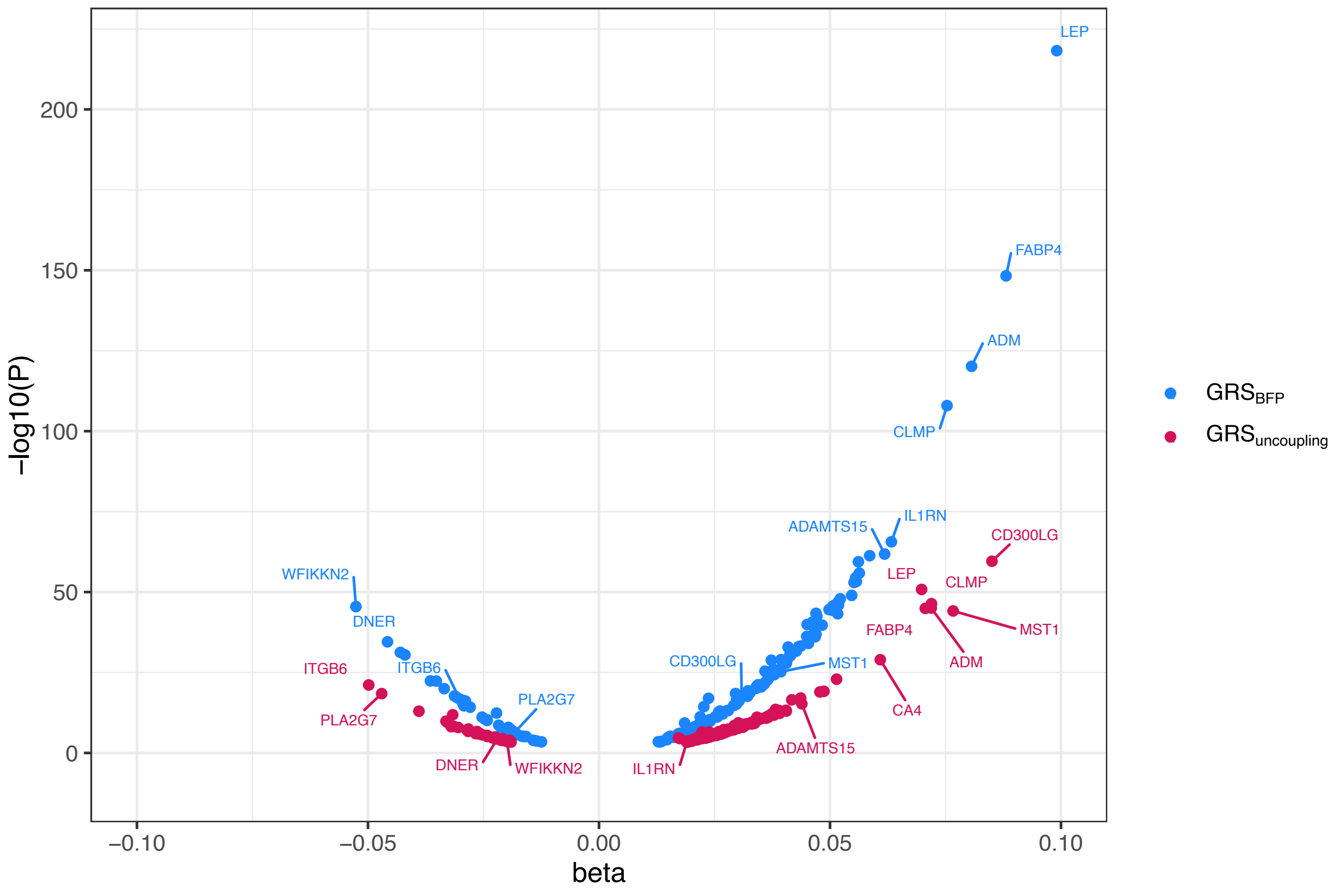

**Extended Data Fig. 6: Plasma proteins (n=129) which associate with the uncoupling- but not the body fat percentage genetic risk score in the UK Biobank.**

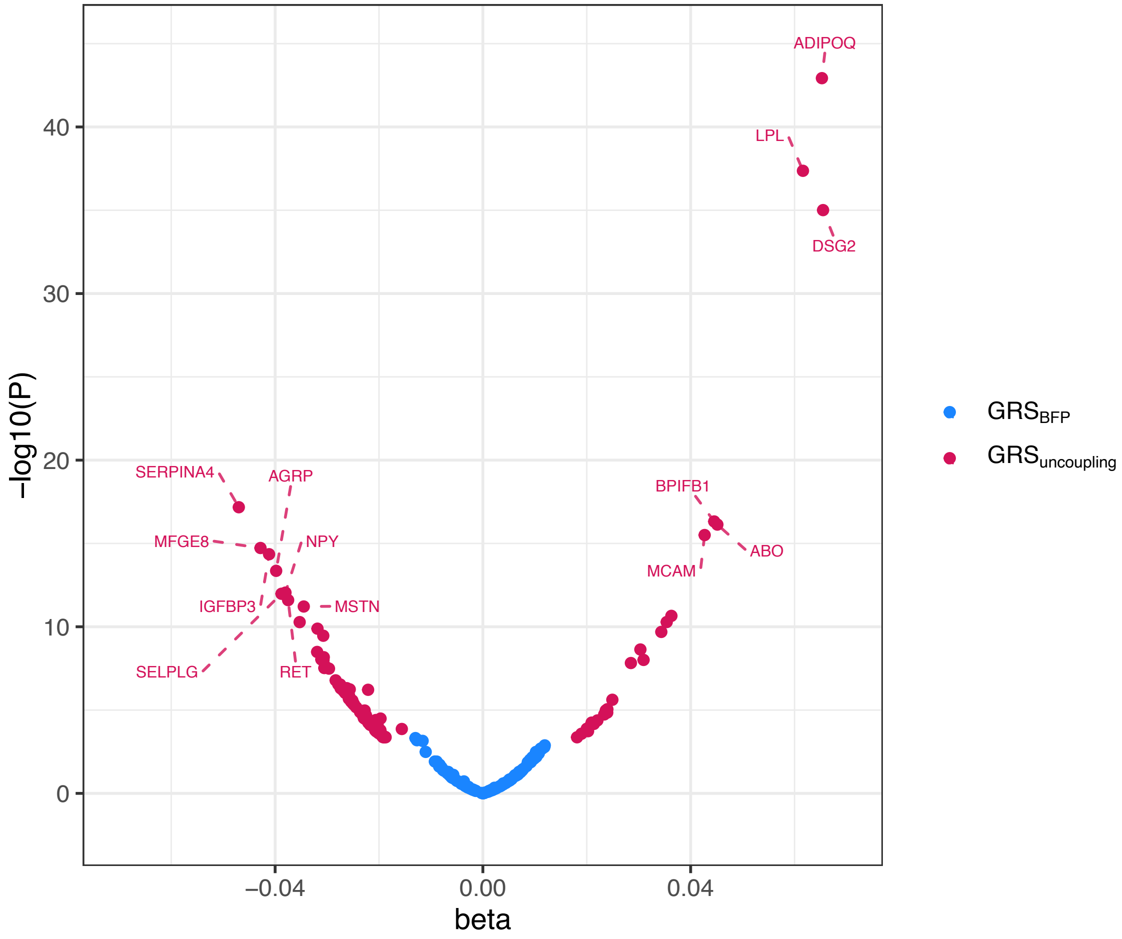
